# Through the patients’ eyes - Psychometric evaluation of the 64-item version of the Experienced Patient-Centeredness Questionnaire (EPAT-64)

**DOI:** 10.1101/2024.03.28.24304955

**Authors:** Eva Christalle, Stefan Zeh, Hannah Führes, Alica Schellhorn, Pola Hahlweg, Jördis Zill, Martin Härter, Carsten Bokemeyer, Jürgen Gallinat, Christoffer Gebhardt, Christina Magnussen, Volkmar Müller, Katharina Schmalstieg-Bahr, André Strahl, Levente Kriston, Isabelle Scholl

**Author notes:** Corresponding author information: Prof. Dr. Isabelle Scholl, Dipl.-Psych., Martinistr. 52, 20246 Hamburg, Germany. Levente Kriston and Isabelle Scholl contributed equally.

## Abstract

**Background:** Patient-reported experience measures (PREMs) are valuable tools for evaluating patient-centeredness (PC) from the patients’ perspective. Despite their utility, a comprehensive PREM addressing PC has been lacking. To bridge this gap, we developed the preliminary version of the Experienced Patient-Centeredness Questionnaire (EPAT), a disease-generic tool based on the integrative model of PC comprising 16 dimensions. It demonstrated content validity. The aim of this study was to test its psychometric properties and to develop a final 64-items version (EPAT-64).

**Methods:** In this cross-sectional study, we included adult patients treated for cardiovascular diseases, cancer, musculoskeletal diseases, and mental disorders in inpatient or outpatient settings in Germany. For each dimension of PC, we selected four items based on item characteristics such as item difficulty and corrected item-total correlation. We tested structural validity using confirmatory factor analysis, examined reliability by McDondald’s Omega, and tested construct validity by examining correlations with general health status and satisfaction with care.

**Results:** Analysis of data from 2.024 patients showed excellent acceptance and acceptable item-total-correlations for all EPAT-64 items, with few items demonstrating ceiling effects. The confirmatory factor analysis indicated the best fit of a bifactor model, where each item loaded on both a general factor and a dimension-specific factor. Omega showed high reliability for the general factor while varying for specific dimensions. Construct validity was confirmed by absence of strong correlations with general health status and a strong correlation of the general factor with satisfaction with care.

**Conclusions:** The EPAT-64 demonstrated commendable psychometric properties. This tool allows comprehensive assessment of PC, offering flexibility to users who can measure each dimension with a 4-item module or choose modules based on their needs. The EPAT-64 serves multiple purposes, including quality improvement initiatives and evaluation of interventions aiming to enhance PC. Its versatility empowers users in diverse healthcare settings.

**What is already known on this topic:** Patient-reported experience measures (PREMs) can be used to assess patient-centeredness (PC) from the patients’ perspective. The “Experienced Patient-Centeredness Questionnaire” (EPAT) is the first PREM to comprehensively assess 16 dimensions of PC.

**What this study adds:** In this study, we tested the psychometric properties of all items developed for the EPAT and developed the 64-item version of the EPAT (EPAT-64), which demonstrated good psychometric properties.

**How this study might affect research, practice or policy:** The EPAT-64 can be used in research and routine care, e.g. to evaluate interventions, provide feedback to healthcare professionals, support quality improvement, set benchmarks, and, consequently improve PC.

## BACKGROUND

Patient-centeredness (PC) is defined as one of the key elements of high quality healthcare worldwide.^1, 2^ It is positively associated with health-related outcomes, such as patient satisfaction, higher knowledge, faster recovery, and better health behaviour.^3–6^ In Germany, as in many other countries^7^, health policy,^8–10^ research,^11–14^ and medical education^15^ have endorsed PC. To overcome ambiguities in the definition of PC, Scholl et al. developed the integrative model of PC,^16^ which was evaluated and extended in two Delphi studies.^17,18^ The model includes 16 dimensions: (1) essential characteristics of the clinician, (2) clinician-patient relationship, (3) patient as a unique person, (4) biopsychosocial perspective, (5) clinician-patient communication, (6) integration of medical and non-medical care, (7) teamwork and teambuilding, (8) access to care, (9) coordination and continuity of care, (10) patient safety, (11) patient information, (12) patient involvement in care, (13) involvement of family and friends, (14) patient empowerment, (15) physical support, and (16) emotional support.

The effective management of PC relies on sound measurement to assess the effects of interventions.^19,20^ For such measurement, it is important to consider the target group of PC, i.e., the patients as recipients of healthcare, e.g. by using patient-reported experience measures (PREMs). PREMs capture patients’ reports directly on whether or how often they have experienced specific processes or behaviours in their healthcare.^21^ This allows the users to derive follow-up actions from the results, such as specific interventions for healthcare professionals or patients. Results from PREMs can be used to assess the quality of healthcare.^22^ Positive patient experiences are associated with patient safety and clinical effectiveness^23^, adherence, better clinical outcomes, and lower rates of healthcare utilization.^24^ A range of PREMs have been developed already.^21^ ^25^ While some of them assess only selective aspects of PC, there is no PREM that considers all 16 dimensions of PC as defined by Scholl and colleagues.^16^ In particular, a recent systematic review by Mihaljevic and colleagues^26^ showed that, to date, there is no German PREM that comprehensively assesses PC.

To bridge this research gap, we developed a preliminary version of the “Experienced Patient-Centeredness Questionnaire” (EPAT), a PREM that assesses PC from the perspective of individuals receiving care^27^. It was developed using a multi-step process including item generation based on a literature review, focus groups with patients, and key informant interviews with experts as well as item selection based on a relevance rating with experts and cognitive interviews with patients. The thorough development process, combining theory-driven approaches and the target groups’ perspective, ensured high content validity of the EPAT. It was developed as a disease-generic questionnaire in German language with an outpatient and an inpatient version. Further, a reflective measurement model was used (i.e., we assumed that specific aspects of patients experience are causally determined by underlying latent dimensions of patient-centeredness). The preliminary version of the EPAT comprised 120 items for outpatients and 121 items for inpatients. Full details of item development and selection process are reported in Christalle et al.^27^ So far, the preliminary version of the EPAT was not psychometrically tested. In addition, a questionnaire of this length is impractical as it consumes lots of resources and is tedious for respondents. Considering that the minimum number of items to estimate and test measurement models is four, a reduction to 64 items (four items for each of the 16 dimensions) would improve its feasibility.

Hence, in the present study, we psychometrically tested the preliminary version of the EPAT. Further, we report on the item selection process for a final version consisting of 64 items (EPAT-64) and its psychometric properties.

## METHODS

### Study design

The development and psychometric testing of the EPAT was part of the ASPIRED study (Assessment of Patient-Centeredness Through Patient-Reported Experience Measures), a 6-year mixed-methods study aiming to assess the patients’ perspective on PC.^27^ ^28^ Here, we report on the psychometric testing of the EPAT. First, the preliminary version of the EPAT was psychometrically tested and item characteristics were calculated. Second, we used those results to select items for the EPAT-64. Third, we assessed structural validity, construct validity, and reliability of the EPAT-64. As there are no guidelines for reporting psychometric evaluations for PREMs, we used the COSMIN guideline on reporting patient-reported outcome measures (PROMs) where applicable (see Appendix 1).^29^

### The EPAT questionnaire

For psychometric testing we used the preliminary version of the EPAT. It comprises of 16 domains.^16^ ^18^ and has two versions, one for inpatients and one for outpatients. All items are rated on a 6-point-Likert-scale of agreement (1 = completely disagree, 2 = strongly disagree, 3 = somewhat disagree, 4 = somewhat agree, 5 = strongly agree, 6 = completely agree). All items had an option to choose ‘does not concern me’. For most items, a high score implied high experienced PC, while 9 items were reversed. The EPAT was developed and psychometrically tested in German. The final version (EPAT-64) was translated into English using a team-based translation process consisting of translation, review, adjudication, pretesting and documentation (TRAPD).^30^

### Data collection and participants

We recruited participants from June 2020 to February 2022. As planned in the protocol^28^, we started recruitment in inpatient and outpatient healthcare institutions (e.g. hospitals, primary care centres) in the metropolitan area of Hamburg, Germany, using consecutive sampling. We used paper-and-pencil questionnaires. Outpatients received the questionnaire on the day of their appointment and were asked to report on their experience during the last four weeks within the respective outpatient clinic. Inpatients received the questionnaire on day of discharge or the day before and were instructed to fill in the questionnaire after discharge and to report on their whole stay. As the recruitment was slowed down by the ongoing COVID-19 pandemic, we increased the recruitment efforts by adding an online survey using LimeSurvey (LimeSurvey GmbH, Hamburg, Germany) starting in December 2020. We recruited participants of the online survey via community-based strategies (i.e., social media, postings in supermarkets, and via notes shared by self-help groups and patient organizations). They were instructed to refer to their last outpatient visit or inpatient stay that should have been no longer than one months ago. Inclusion criteria were age of 18 years or older and being currently treated for at least one disease from the following four disease groups: cardiovascular diseases, cancer, musculoskeletal diseases, and mental disorders. Inclusion was based on self-report. We aimed to include 250 inpatients and 250 outpatients for each of the four disease groups, respectively, resulting in a total target sample size of 2000.^28^

In addition to the preliminary version of the EPAT, participants were also asked to complete the first item of the German version of the 12-item Short Form Survey, which assesses general health status.^31^ This item has an excellent item difficulty of 0.4 and high acceptability (less than 4 % missing values). We assessed satisfaction with care with the German version of the 8-item Client Satisfaction Questionnaire (ZUF-8),^32^ a unidimensional questionnaire with high internal consistency (Cronbach’s alpha = 0.9). We also administered the European Health Literacy Questionnaire (HLS-EU-Q16),^33^ a short version of the HLS-EU-Q47 with low ceiling effects and strong correlation with the long version (r = 0.82). Furthermore, we included questions on demographic factors (e.g., gender and age) and health status (e.g., years since receiving the corresponding diagnosis). Finally, two control items were introduced first in the online version and starting from June 2021 in the paper-pencil-version (‘To show that you are reading attentively, please check “somewhat disagree”/”somewhat agree”.’). Data collection was anonymous. All participants gave written consent. Paper questionnaires were sent back with a return envelope free of charge. Participants had the possibility to receive an incentive of 10€ upon completion of the questionnaire.

### Data analyses

First, we analysed all items of the preliminary version. Then, we selected items for the EPAT-64 in a team discussion with five members (IS, LK, MH, JZ, EC). All members are experts in the field of PC. Three of them (IS, JZ, EC) were part of the item development process and were familiar with the qualitative data that the items were based on.^27^ For each dimension, we chose four items resulting in 64 items covering 16 dimensions of PC. We considered the following psychometric properties during item selection: (1) percentage of missing values as proxy for acceptance, (2) percentage of patients replying ‘does not concern me’ as proxy for relevance, (3) item-difficulty calculated by standardising the mean to range from 0 to 1 (recommended range 0.2 to 0.8),^34^ (4) within-dimension inter-item-correlations (recommended range >0.3)^35^, (5) within-dimension corrected item-total correlation (recommended range >0.3),^35^ and (6) content validity (e.g., how important an item is to the definition of the given dimension, how often an item showed up in the qualitative data used for item development, or to which degree it covers a different aspect than other selected items).

After having selected the 64 items for the EPAT-64, we examined structural validity by confirmatory factor analysis (CFA) using a robust version of maximum likelihood estimator (MLR).^36^ Based on our theory, we tested five different models: (1) unidimensional model, (2) correlated first-order dimension model, (3) hierarchical model, (4) bifactor model with uncorrelated dimensions, and (5) bifactor model with correlated dimensions. For detailed descriptions and depictions of all models, please refer to Appendix 2. For examination of model fit and selection, we followed recommendations by using a combination of model fit indices:^37^ Root Mean Square Error of Approximation (RMSEA), Standardized Root Mean Square Residual (SRMR), Tucker-Lewis Index (TLI), Comparative Fit Index (CFI). For comparison of models we used Akaike information criterion (AIC) and Bayesian information criterion (BIC). Further, we report the chi-squared test statistic of model fit.^38^ For the selected model, we report standardised factor loadings. We planned to test this model in the four disease groups to examine measure invariance between groups.^35^ Further, we examined reliability by McDonald’s Omega hierarchical which allows to estimate reliability in models with a bifactor structure.^39^

Construct validity was assessed by correlating the model-based factor scores of each dimension with two other measures as described in the study protocol.^28^ First, discriminant validity was tested by examining the correlations with general health status, measured by the first item of the German version of 12-Item Short Form Survey.^31^ We hypothesized, that the magnitude of these correlations should be below 0.3.^40^ Second, for convergent validity, we used correlations with satisfaction with care measured by the German version of the 8-item Client Satisfaction Questionnaire (ZUF-8).^32^ Here, we expected the correlations to exceed 0.5.^40^

We used SPSS Version 27.0 (IBM Corp., Armonk, NY, USA) to enter data from the paper-and-pencil version, to check their validity by a second person, to import data from LimeSurvey, and to calculate sample characteristics. All other data cleaning and analyses were done with R Version 4.3.2 (R Core Team, Vienna, Austria). We excluded participants if they responded to less than 70 % of EPAT questions or if they failed both control items. Where applicable, we used full information maximum likelihood to deal with missing values.^41^ We analysed data separately for inpatient and outpatient settings. Here, we report the results for all four disease groups combined.

## RESULTS

### Sample

For the paper-and-pencil version, we asked 4,788 outpatients if they wanted to participate, of whom 2,357 gave consent and 905 returned the questionnaire. Further, we asked 3,046 inpatients, of whom 1,799 gave their consent and 704 returned the questionnaire. For the online questionnaire, 1,042 patients consented. We have no information on patients that did not consent or did not finish the questionnaire. Hence, we were not able to perform dropout analyses. In total, questionnaires from 1,092 outpatients and 931 inpatients were included in our analyses (see flowchart in Figure 1).

**Figure 1:**
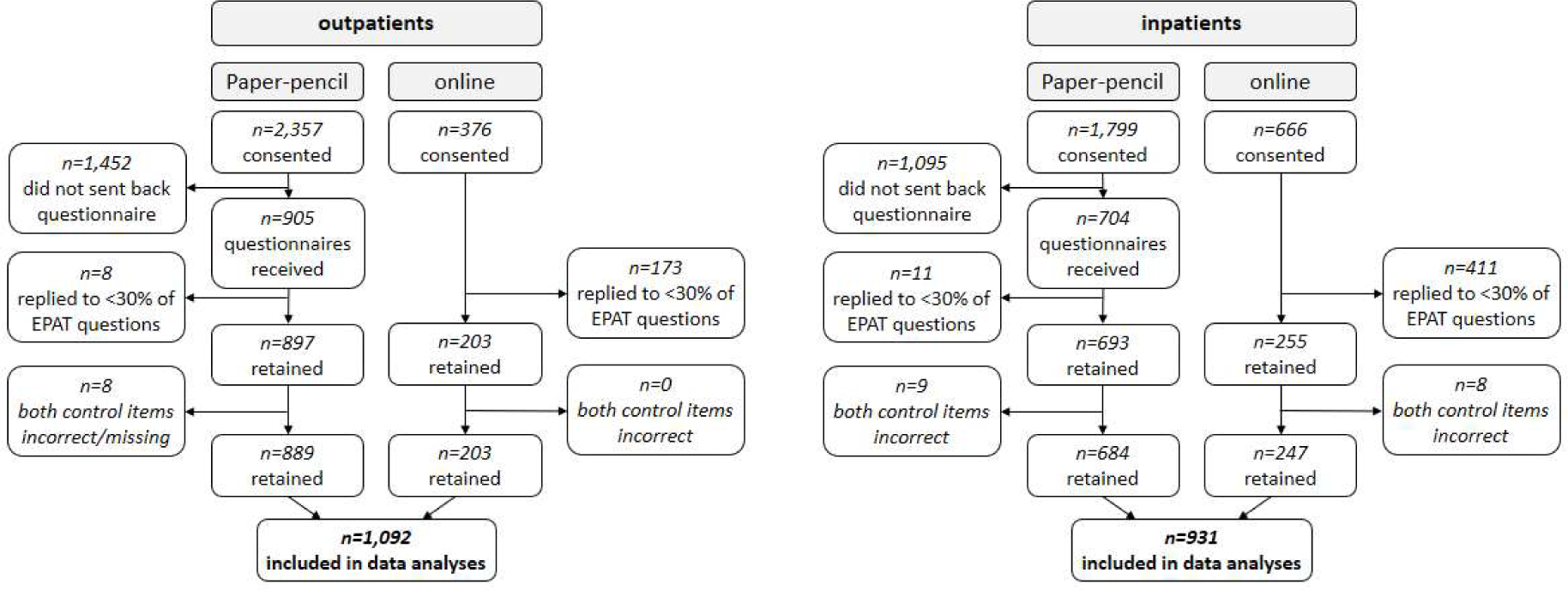
Participant flow chart.

Of the outpatients included in psychometric analyses, 59.1 % were female, 38.5 % male, and 0.5 % chose the diverse option (e.g. trans, inter, non-binary). Their mean age was 52.8 years (standard deviation (SD) = 17.4, range 18-95) and mean time since they were first diagnosed with the respective disease was 11.4 years (SD = 11.0). Of the inpatients, 41.3 % were female, 55.5 % male, and 0.6 % identified as diverse. Their mean age was 55.5 years (SD = 17.4, range 18-92) and mean time since the respective diagnosis was 8.5 years (SD = 10.0). Further details on demographic and health-related variables are shown in Table 1. For subgroup sample characteristics refer to Appendix 3.

**Table 1:**
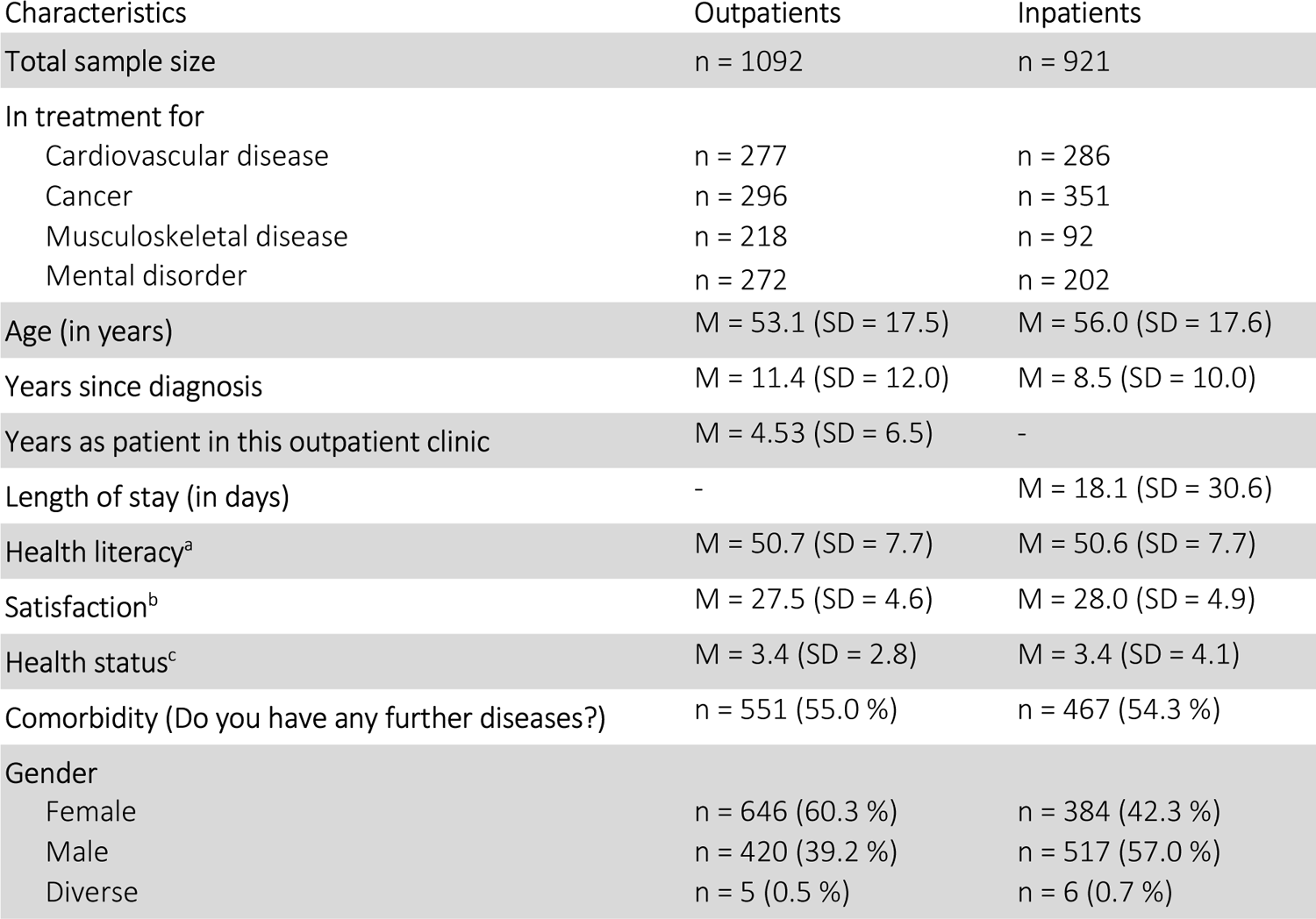
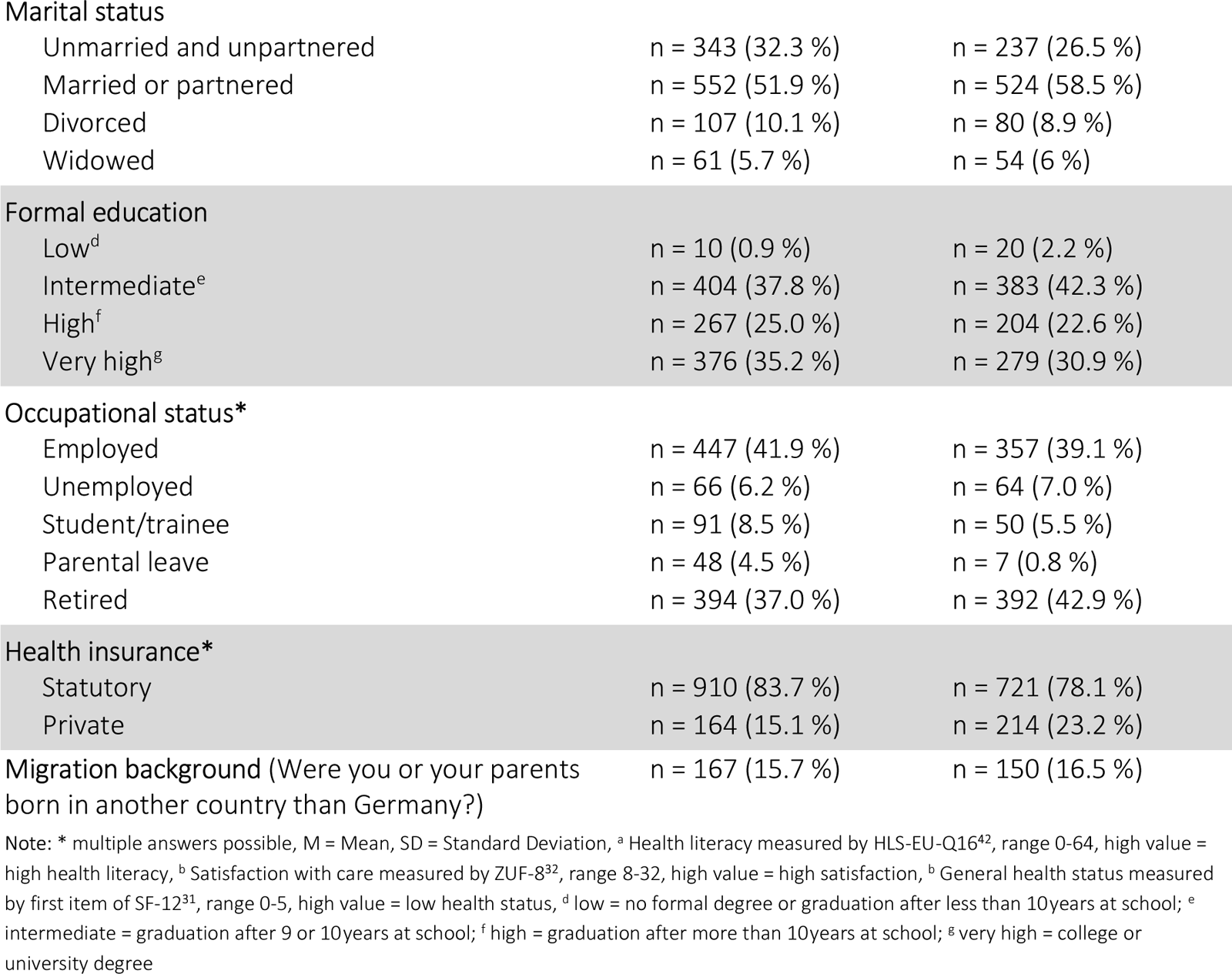
Sample characteristics.

| Characteristics | Outpatients | Inpatients |
| --- | --- | --- |
| **Total sample size** | n = 1092 | n = 921 |
| **In treatment for** |  |  |
| Cardiovascular disease | n = 277 | n = 286 |
| Cancer | n = 296 | n = 351 |
| Musculoskeletal disease | n = 218 | n = 92 |
| Mental disorder | n = 272 | n = 202 |
| **Age (in years)** | M = 53.1 (SD = 17.5) | M = 56.0 (SD = 17.6) |
| **Years since diagnosis** | M = 11.4 (SD = 12.0) | M = 8.5 (SD = 10.0) |
| **Years as patient in this outpatient clinic** | M = 4.53 (SD = 6.5) | - |
| **Length of stay (in days)** | - | M = 18.1 (SD = 30.6) |
| **Health literacy<sup>a</sup>** | M = 50.7 (SD = 7.7) | M = 50.6 (SD = 7.7) |
| **Satisfaction<sup>b</sup>** | M = 27.5 (SD = 4.6) | M = 28.0 (SD = 4.9) |
| **Health status<sup>c</sup>** | M = 3.4 (SD = 2.8) | M = 3.4 (SD = 4.1) |
| **Comorbidity (Do you have any further diseases?)** | n = 551 (55.0 %) | n = 467 (54.3 %) |
| **Gender** |  |  |
| Female | n = 646 (60.3 %) | n = 384 (42.3 %) |
| Male | n = 420 (39.2 %) | n = 517 (57.0 %) |
| Diverse | n = 5 (0.5 %) | n = 6 (0.7 %) |
| **Marital status** |  |  |
| Unmarried and unpartnered | n = 343 (32.3 %) | n = 237 (26.5 %) |
| Married or partnered | n = 552 (51.9 %) | n = 524 (58.5 %) |
| Divorced | n = 107 (10.1 %) | n = 80 (8.9 %) |
| Widowed | n = 61 (5.7 %) | n = 54 (6 %) |
| **Formal education** |  |  |
| Low<sup>d</sup> | n = 10 (0.9 %) | n = 20 (2.2 %) |
| Intermediate<sup>e</sup> | n = 404 (37.8 %) | n = 383 (42.3 %) |
| High<sup>f</sup> | n = 267 (25.0 %) | n = 204 (22.6 %) |
| Very high<sup>g</sup> | n = 376 (35.2 %) | n = 279 (30.9 %) |
| **Occupational status\*** |  |  |
| Employed | n = 447 (41.9 %) | n = 357 (39.1 %) |
| Unemployed | n = 66 (6.2 %) | n = 64 (7.0 %) |
| Student/trainee | n = 91 (8.5 %) | n = 50 (5.5 %) |
| Parental leave | n = 48 (4.5 %) | n = 7 (0.8 %) |
| Retired | n = 394 (37.0 %) | n = 392 (42.9 %) |
| **Health insurance\*** |  |  |
| Statutory | n = 910 (83.7 %) | n = 721 (78.1 %) |
| Private | n = 164 (15.1 %) | n = 214 (23.2 %) |

**Note:** \* multiple answers possible, M = Mean, SD = Standard Deviation, <sup>a</sup> Health literacy measured by HLS-EU-Q16<sup>42</sup>, range 0-64, high value = high health literacy, <sup>b</sup> Satisfaction with care measured by ZUF-8<sup>32</sup>, range 8-32, high value = high satisfaction, <sup>c</sup> General health status measured by first item of SF-12<sup>31</sup>, range 0-5, high value = low health status, <sup>d</sup> low = no formal degree or graduation after less than 10years at school; <sup>e</sup> intermediate = graduation after 9 or 10years at school; <sup>f</sup> high = graduation after more than 10years at school; <sup>g</sup> very high = college or university degree

### Item characteristics

For outpatients, all 64 selected items showed high acceptability (highest percentage of missing responses was 2.7 %). Regarding relevance, 47 items had less than 30 % of participants answering ‘does not concern me’ and seven items had a rate over 50 %. While 46 items showed excellent item difficulty (between 0.2 and 0.8), 18 items showed ceiling effects (item difficulty >0.8). All but one item had an item-total correlation greater than 0.3.

For inpatients, again, all 64 items had high acceptability (highest percentage of missing responses was 2.7 %). Regarding relevance, we included 55 items with a ‘does not concern me’ rate below 30 % and two items above 50 %. Item difficulty was excellent for 42 items, and 22 items showed ceiling effects. All items had item-total correlation greater than 0.3.

Item characteristics of all tested items are shown in Table 2. For item wording, please refer to the full questionnaires at www.uke.de/epat.

**Table 2:**
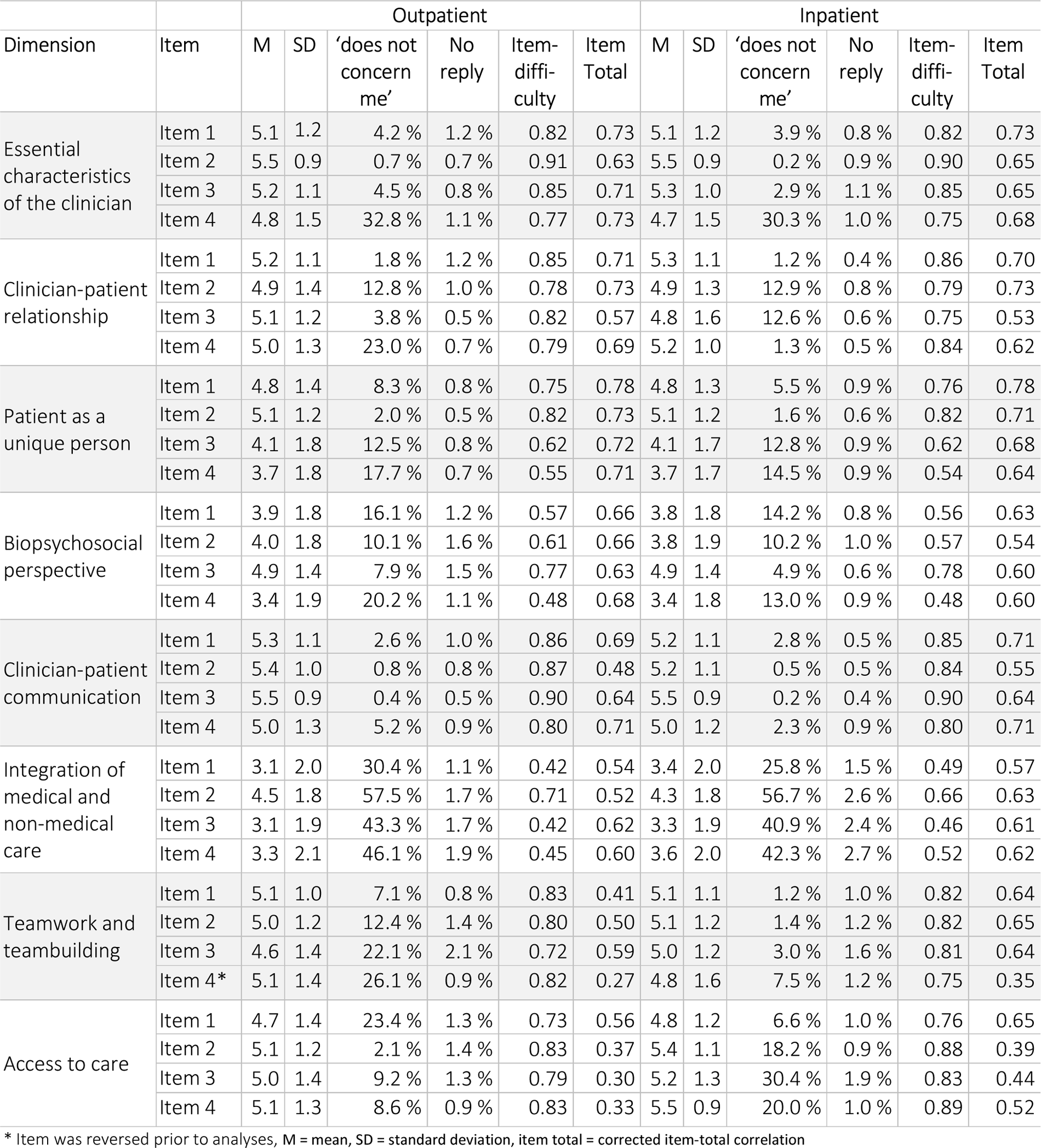
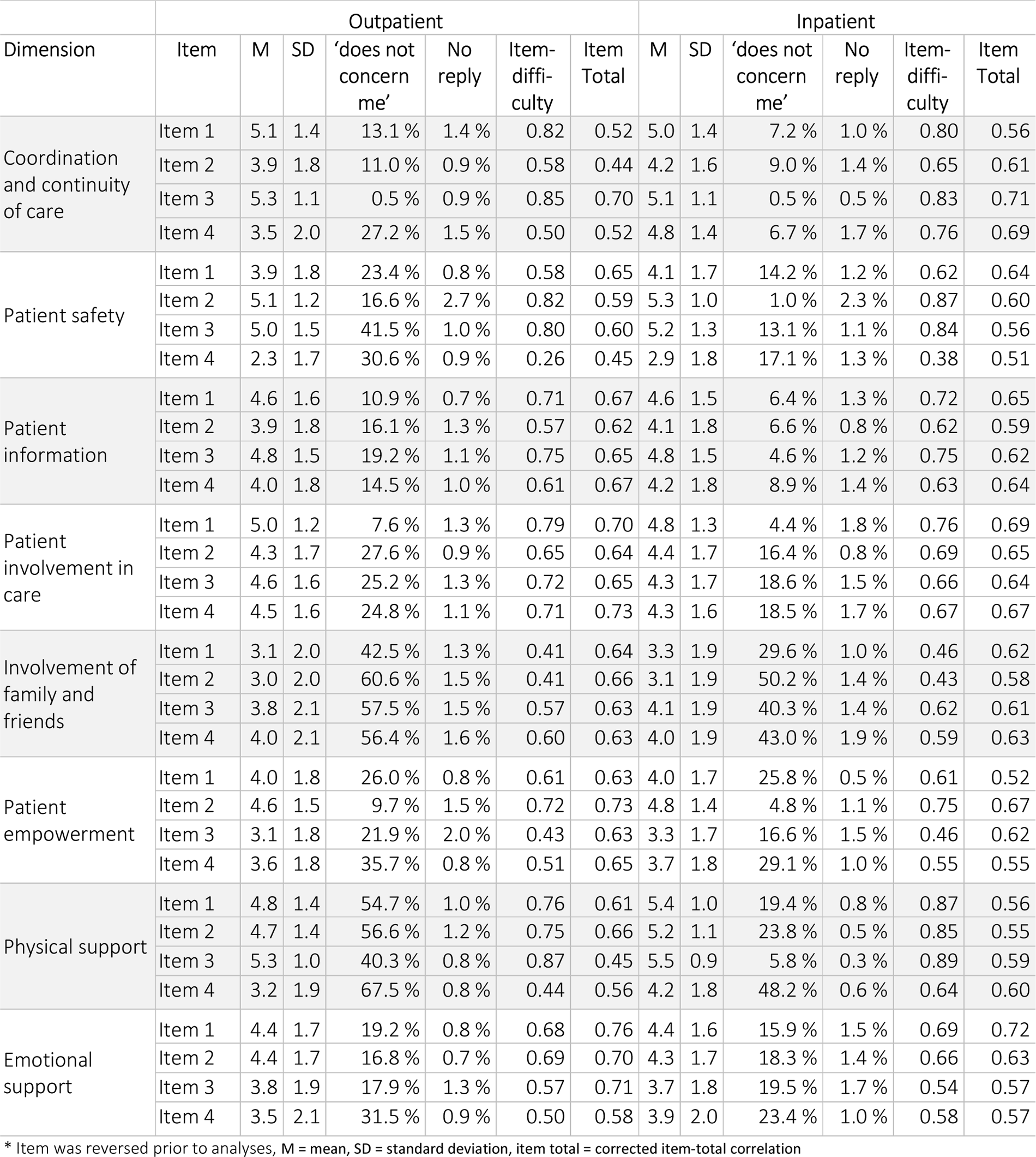
Items characteristics of the EPAT-64.

Item characteristics per subgroup are shown in Appendix 4 (outpatient sample) and Appendix 5 (inpatient sample). Item characteristics for all tested items can be requested from the authors.

### Structural validity

The model fit indices for the five tested models are shown in Table 3.

**Table 3:** Model fit indices for confirmatory factor analysis on EPAT-64.

|  |  | Chi square |  |  | RMSEA | SRMR | TLI | CFI | AIC | BIC |
| --- | --- | --- | --- | --- | --- | --- | --- | --- | --- | --- |
| | | $\chi^2$ | df | P value | | | | | | |
| Recommendation |  |  |  | >.05 | ≤ .06 | <.08 | ≥ .90 | ≥ .90 | Model with smallest value |  |
| Outpatient version | 1 Unidimensional | 14957 | 1952 | <.001 | .078 | .087 | .659 | .670 | 166616 | 167575 |
|  | 2 Correlated first-order | 7098 | 1832 | <.001 | .051 | .072 | .853 | .866 | 158996 | 160555 |
|  | 3 Hierarchical | 8556 | 1936 | <.001 | .056 | .084 | .825 | .832 | 160247 | 161286 |
|  | 4 Bifactor without dimension correlations | No results. Model estimation did not converge. |  |  |  |  |  |  |  |  |
|  | 5 Bifactor with dimension correlations | 4871 | 1768 | <.001 | .040 | .055 | .910 | .921 | 156898 | 158776 |
| Inpatient version | 1 Unidimensional | 15201 | 1952 | <.001 | .085 | .101 | .631 | .643 | 151891 | 152819 |
|  | 2 Correlated first-order | 7276 | 1832 | <.001 | .056 | .083 | .838 | .853 | 144206 | 145714 |
|  | 3 Hierarchical | 8839 | 1936 | <.001 | .062 | .102 | .806 | .814 | 145561 | 146567 |
|  | 4 Bifactor without dimension correlations | No results. Model estimation did not converge. |  |  |  |  |  |  |  |  |
|  | 5 Bifactor with dimension correlations | 4721 | 1768 | <.001 | .042 | .051 | .909 | .920 | 141779 | 143597 |

The best fit was found for the bifactor model with correlated dimensions. Here, all items load on their respective specific dimension as well as a general dimension. Specific dimensions are intercorrelated but their correlation with the general factor was restricted to be zero. Standardised factor loadings of this model are reported in Table 5. Correlations between specific dimension showed a wide variability ranging from -0.140 to 0.824 (see Appendix 6).

**Table 5:** Standardised actor loadings and reliability coefficients of the correlated bifactor model.

|  | Dimension | Loadings on general factor |  |  |  | Loadings on specific dimension |  |  |  | Reliability |
| --- | --- | --- | --- | --- | --- | --- | --- | --- | --- | --- |
|  |  | Item 1 | Item 2 | Item 3 | Item 4 | Item 1 | Item 2 | Item 3 | Item 4 | Omega_H |
| Outpatient sample | Essential characteristics of the clinicians | 0.759 | 0.795 | 0.767 | 0.575 | 0.440 | 0.216 | 0.387 | 0.571 | 0.214 |
|  | Clinician-patient relationship | 0.754 | 0.682 | 0.565 | 0.627 | 0.474 | 0.595 | 0.225 | 0.345 | 0.242 |
|  | Patient as a unique person | 0.704 | 0.795 | 0.366 | 0.288 | 0.498 | 0.332 | 0.782 | 0.833 | 0.515 |
|  | Biopsychosocial perspective | 0.495 | 0.290 | 0.352 | 0.243 | 0.488 | 0.805 | 0.717 | 0.814 | 0.705 |
|  | Clinician-patient communication | 0.760 | 0.457 | 0.739 | 0.608 | 0.371 | 0.424 | 0.384 | 0.498 | 0.256 |
|  | Integration of medical and non-medical care | 0.143 | 0.218 | 0.188 | 0.165 | 0.836 | 0.731 | 0.829 | 0.854 | 0.859 |
|  | Teamwork and teambuilding | 0.347 | 0.417 | 0.493 | 0.390 | 0.576 | 0.689 | 0.518 | 0.884 | 0.415 |
|  | Access to care | 0.475 | 0.323 | 0.235 | 0.283 | 0.502 | 0.712 | 0.638 | 0.535 | 0.599 |
|  | Coordination and continuity of care | 0.430 | 0.198 | 0.739 | 0.291 | 0.345 | 0.516 | 0.318 | 0.505 | 0.355 |
|  | Patient safety | 0.458 | 0.359 | 0.586 | 0.114 | 0.453 | 0.683 | 0.298 | 0.685 | 0.506 |
|  | Patient information | 0.489 | 0.374 | 0.534 | 0.426 | 0.620 | 0.649 | 0.564 | 0.665 | 0.559 |
|  | Patient involvement in care | 0.732 | 0.447 | 0.506 | 0.529 | 0.356 | 0.566 | 0.651 | 0.734 | 0.461 |
|  | Involvement of family and friends | 0.187 | 0.201 | 0.233 | 0.255 | 0.882 | 0.895 | 0.825 | 0.791 | 0.872 |
|  | Patient empowerment | 0.232 | 0.570 | 0.227 | 0.224 | 0.764 | 0.515 | 0.732 | 0.790 | 0.718 |
|  | Physical support | 0.567 | 0.586 | 0.457 | 0.190 | 0.706 | 0.725 | 0.209 | 0.443 | 0.461 |
|  | Emotional support | 0.516 | 0.464 | 0.359 | 0.196 | 0.755 | 0.766 | 0.857 | 0.680 | 0.736 |
| General factor |  |  |  |  |  |  |  |  |  | 0.868 |
| Inpatient sample | Essential characteristics of the clinicians | 0.783 | 0.830 | 0.758 | 0.575 | 0.312 | 0.155 | 0.230 | 0.517 | 0.128 |
|  | Clinician-patient relationship | 0.785 | 0.674 | 0.323 | 0.657 | 0.338 | 0.539 | 0.612 | 0.263 | 0.291 |
|  | Patient as a unique person | 0.720 | 0.775 | 0.340 | 0.250 | 0.444 | 0.238 | 0.779 | 0.869 | 0.484 |
|  | Biopsychosocial perspective | 0.257 | 0.180 | 0.445 | 0.263 | 0.813 | 0.776 | 0.476 | 0.826 | 0.757 |
|  | Clinician-patient communication | 0.744 | 0.559 | 0.727 | 0.798 | 0.334 | 0.483 | 0.352 | 0.395 | 0.215 |
|  | Integration of medical and non-medical care | 0.233 | 0.286 | 0.265 | 0.252 | 0.833 | 0.835 | 0.824 | 0.846 | 0.847 |
|  | Teamwork and teambuilding | 0.718 | 0.744 | 0.643 | 0.448 | 0.465 | 0.425 | 0.553 | 0.918 | 0.213 |
|  | Access to care | 0.692 | 0.541 | 0.528 | 0.655 | 0.114 | 0.538 | 0.711 | 0.821 | 0.217 |
|  | Coordination and continuity of care | 0.521 | 0.459 | 0.784 | 0.659 | 0.288 | 0.518 | 0.233 | 0.355 | 0.199 |
|  | Patient safety | 0.392 | 0.734 | 0.558 | 0.256 | 0.695 | 0.123 | 0.249 | 0.726 | 0.365 |
|  | Patient information | 0.489 | 0.378 | 0.592 | 0.538 | 0.622 | 0.673 | 0.443 | 0.575 | 0.493 |
|  | Patient involvement in care | 0.695 | 0.534 | 0.532 | 0.479 | 0.378 | 0.564 | 0.676 | 0.738 | 0.474 |
|  | Involvement of family and friends | 0.273 | 0.265 | 0.415 | 0.378 | 0.850 | 0.862 | 0.689 | 0.755 | 0.780 |
|  | Patient empowerment | 0.212 | 0.572 | 0.330 | 0.223 | 0.742 | 0.398 | 0.687 | 0.866 | 0.668 |
|  | Physical support | 0.662 | 0.670 | 0.677 | 0.415 | 0.622 | 0.632 | 0.169 | 0.230 | 0.268 |
|  | Emotional support | 0.493 | 0.327 | 0.186 | 0.272 | 0.735 | 0.846 | 0.878 | 0.619 | 0.768 |
| General factor |  |  |  |  |  |  |  |  |  | 0.908 |

We further explored the general factor in the bifactor model by calculating correlations with other measures. We found a strong correlation between the model-based scores of the general factor and satisfaction with care as measured by the overall score of ZUF-8 (0.73 for the outpatient sample and 0.83 for the inpatient sample). The magnitude of correlations between the general factor and health literacy, general health status, comorbidity, age, and time since first diagnosis of the corresponding disease, respectively, were below 0.3.

We were not able to examine measurement invariance between different disease groups. The combination of small sample sizes within the groups, high rates of missing values due to the “does not concern me” option, and the complexity of the tested model with a high number of parameters to be estimated led to estimation problems (e.g., models did not converge or the variance-covariance matrix of the estimated parameters was not positive definite). This problem persisted when we used multiple imputation to impute missing data or when we excluded the three dimensions with the highest rate of “does not concern me” replies.

### Reliability

McDonald’s Omega hierarchical was high for the general factor (0.868 for outpatients, 0.908 for inpatients) and varied widely for the specific dimensions (range 0.214 to 0.872 for outpatients and 0.128 to 0.847 for inpatients). In order to support interpretability of sum scores for specific dimensions, we examined their correlations with the model-based factor scores. The results suggest that the sum scores measure a mixture of the general factor and the specific dimensions. The median correlation of the dimension specific sum scores with the general factor score estimate was 0.558 in the outpatient sample and 0.639 in the inpatient sample. The median correlation with the factor score of their corresponding dimension was 0.811 in the outpatient sample and 0.745 in the inpatient sample, respectively (Appendix 7). These coefficients suggest that the proportion of explained variance attributable to the general factor in subscale scores ranges approximately between one and three thirds for most dimensions.

### Construct validity

Using model-based factor scores, discriminant validity was confirmed for all dimensions and the general factor, that is the magnitude of all correlations with general health status were below 0.3. Convergent validity was confirmed for the general factor, which was strongly correlated with satisfaction with care, while the specific dimensions showed no strong correlations. Correlations for all dimensions and the general factor are shown in Appendix 8.

## DISCUSSION

### Summary of the findings

The EPAT-64 is a PREM to assess 16 dimensions of PC from the patients’ perspective. All items demonstrated high acceptability. Most items had low rates of patients answering ‘does not concern me’. Regarding item difficulty, most items were within the recommended range, yet a few items showed ceiling effects. Item-total correlations were good for all but one item. CFA showed the best fit for a bifactor model. In terms of construct validity, discriminant validity was confirmed for all dimensions and the general factor. Convergent validity was confirmed for the general factor. McDonalds Omega hierarchical suggests good reliability of the general factor, while there was a wide variability for the specific dimensions.

### Strengths and limitations

The EPAT-64 is the first PREM that comprehensively assesses PC.^25^ ^26^ One of its strengths is the thorough development and item selection process, guided by a model derived from literature and input from the target group.^16^ ^18^ ^27^ This was continued during the item selection process after psychometric testing. The results above show that not all items included in the EPAT-64 have excellent psychometric properties, in particular regarding ceiling effects, which may limit the ability to discriminate between high and low PC. These items were included for a number of reasons. We decided to choose four items for each dimension, as we aimed for parsimony but needed at least four items for statistical model identifiability.^43^ For some dimensions, the preliminary version of the EPAT had only four items.^27^ This meant that we had to either include all items or delete the whole dimension. However, as our Delphi study prior to questionnaire development showed that all dimensions were relevant to patients,^18^ deleting dimensions would have reduced the comprehensiveness and hence the content validity of the EPAT-64. Furthermore, we considered not only the quantitative characteristics of the items but also their content. We would like to argue that selecting items solely on the basis of quantitative data risks compromising content validity.^43^ It may lead to more reliable questionnaires with better discrimination between groups, but it also runs the risk of deleting items whose content is essential for measuring the given construct.

The CFA showed best fit for a bifactor model with correlated dimensions. This model postulates that all items load both on a general factor and on their respective specific dimension, which is independent of the general factor. A strength of the bifactor model that is enables users to partition scores into an estimation of the general factor and of the specific dimension.^44^ We suspect that the general factor captures more subjective aspects of PC as it is strongly correlation with satisfaction with care. The variance in the data that is not captured by the general factor seems to be well explained by the postulated dimensions of PC, which is a confirmation of the underlying conceptual model. As these dimensions are independent of the (presumably) subjective general factor, we assume that they capture more objective aspects of PC, i.e., rather if something happened than how well it went. This corresponds to frequent definitions of PREMs^21^. To be able to evaluate both aspects of PC, we strongly recommend users to apply the whole EPAT-64 and model the bifactor model with their own data set. This way, scores can be calculated separately, first for the more subjective general factor, and secondly a more objective score for each dimension. Yet, in routine care this might be infeasible as often there are not sufficient resources to administer and analyse a questionnaire with 64 items or there is a lack of time or statistical expertise to estimate bifactor models. Here, users might want to choose just the most relevant dimensions for their context and calculate sum scores. This approach gives a high degree of flexibility, which allows users to adapt the questionnaire to their question, area of application, and resources. We suggest that sum scores are calculated per dimension only for respondents with one or less missing values. One missing value per dimension can be replaced by the mean of the other three items for each respondent. For respondents with two or more missing values in one dimension, no sum score should be calculated for this dimension. When interpreting those sum scores one should consider that this score combines subjective and objective PC assessments, which cannot be well differentiated based on sum scores alone. Further, the dimensions have different degrees of reliability when estimated without the general factor. For example, “Integration of medical and non-medical care” seems to be more appropriate to be administered as a stand-alone dimension than “Essential characteristics of the clinicians”, based on their reliability estimated by Omega hierarchical. In addition, the correlations between the specific dimensions makes the interpretation of their sum scores more challenging. Appendix 7 can help to guide interpretation of sum scores, but more research is needed.

A limitation is that we were not able to examine measurement invariance between different disease groups. Items were developed generically and we tested the EPAT in four disease groups (cardiovascular, cancer, musculoskeletal, mental health). As these diseases affect a large proportion of the population and show different courses and treatment pathways, we hope that the results will be transferable to other disease groups as well. Yet, due to the complex measurement model and the large number of ‘does not concern me’-replies in some items, we were not able to confirm that the EPAT does indeed have a common factor structure within each subgroup.

### Implications

There is a wide range of possible applications of the EPAT-64. Measuring PC is essential for research on the current state of PC as well as on interventions aiming to foster PC. Internationally, PREMs are used to provide feedback to healthcare providers, inform quality improvement, public reporting, benchmarking, and value-based purchasing.^45–47^ As Coulter and colleagues discussed, measurement alone is not sufficient to improve patient experiences.^48^ The resulting data need to be used for quality improvement in a coordinated approach (including senior leadership, clear goals and continuous performance measurement).^48^ Nevertheless, there is emerging evidence that being informed about patient experiences is associated with improved communication^49^ and meetings to discuss PREM results are associated with improved patient experience.^46^

Psychometric testing in independent samples are indispensable to ensure that the psychometric properties found here hold in other populations and contexts. Particular attention should be paid to the interpretation of the bifactor structure with correlated dimensions and measurement invariance, that is whether the factor structure found here can be replicated for different subgroups. In addition to validity and reliability, aspects of fairness and responsiveness should be investigated.^50^ Regarding testing in other patient groups, the EPAT-64 is currently adapted to assess PC in the context of healthcare and support services for women with unintended pregnancy.^51^ Further adaptations to other healthcare contexts, such as rehabilitation facilities or non-academic hospitals, could also provide useful insights.

Internationally, there is no comparable PREM that comprehensively assesses all dimensions of PC.^25^ Therefore, cross-cultural adaptations and translations of the EPAT-64 would provide an opportunity to promote PC around the world. In addition, translations are helpful to reach parts of the population in Germany that are not fluent in German. Table 2 contains an official translation into English using the TRAPD process.^30^ However, further tests on comprehensibility and psychometric properties are required.

## Conclusion

The EPAT-64, a novel questionnaire assessing 16 dimensions of patient-centeredness from the patient’s perspective, emerges from a meticulous development process that blends theory with data driven decisions. Psychometric testing within a substantial patient cohort underscores its robust psychometric properties. Nevertheless, further research is imperative to refine the interpretation of scale scores. The adaptability of this tool to specific purposes positions it as a valuable instrument for both assessment and cultivation of patient-centeredness.

## Supporting information

Appendix 1 COSMIN reporting guideline

Appendix 2 Confirmatory factor analyses

Appendix 3 Subgroup sample characteristics

Appendix 4 Item characteristics per medical condition - outpatient sample

Appendix 5 Item characteristics per medical condition

Appendix 6 Correlations dimensions

Appendix 7 Correlations sum scores

Appendix 8 Construct validity

## Data Availability

All data produced in the present study are available upon reasonable request to the authors.

https://www.uke.de/epat

## ACKNOWLEDGEMENTS

We thank our student assistant Sibel Gürü and our research interns Lucas Bolz, Antonia Buchal, Sophia Ehrentraut, Carolin Guddat, Zoe Henning, Constanze Mahn, Yannick Mosemann, Philip Niemann, Svea-Marie Nohrden, Paul Packheiser, Ronja Platzeck, Mareike Remmers, Ramona Rusch, Laura Stefan, Melvin Schneider, and Lisa Strelow for carrying out data collection. Further, we would like to thank all our collaboration partners who gave us access to their patients for the data collection: Klinik für Psychosomatische Medizin und Psychotherapie (UKE), III. Medizinische Klinik und Poliklinik (UKE), Strahlentherapie & Radioonkologie (UKE), Martiniklinik, Evangelisches Krankenhaus Alsterdorf, Verhaltenstherapie Falkenried, Bundeswehrkrankenhaus Hamburg, Asklepios Westklinikum Hamburg, Psychiatrische Ambulanz Dr. Guth, and Fachzentrum Altona für Psychiatrie und Psychotherapie.

## AUTHORS CONTRIBUTIONS

IS was the responsible principle investigator of the study. IS, LK and MH were involved in planning and preparation of the study. SZ, AS, HF and EC recruited participants and collected data. EC analysed data with supervision by LK. EC, SZ, JZ, IS, MH, PH and LK interpreted results and discussed the item selection. EC wrote the first draft of the manuscript. CB, JG, CG, CM, VM, KSB and AS acitively supported data collection. All authors critically revised the manuscript for important intellectual content. All authors gave final approval of the version to be published and agreed to be accountable for the work.

## COMPETING INTERESTS

PH declares to have no financial conflicts of interest. PH is a board member of the International Shared Decision Making Society, a charitable scientific society. PH received research funding from German Research Foundation, University of Hamburg and Robert-Bosch-Foundation. IS received honoraria for presentations and speeches on patient-centered care from the following commercial entities: onkowissen.de GmbH, ClinSol GmbH & Co. KG. All other authors declare that they have no competing interests.

## FUNDING STATEMENT

This study was funded by the German Federal Ministry of Education and Research (Bundesministerium für Bildung und Forschung – BMBF) with the grant number 01GY1614. The funder had no role in decision to publish or preparation of the manuscript.

## ETHICS APPROVAL AND CONSENT TO PARTICIPATE

The study was carried out according to the latest version of the Helsinki Declaration of the World Medical Association. Principles of good scientific practice were respected. The study had been approved by the Ethics Committee of the Medical Association Hamburg (study ID: PV5724). Study participation was voluntary and no foreseeable risks for participants resulted from the participation in this study. Participants were fully informed about the aims of the study, data collection, and the use of collected data. Written informed consent was obtained prior to participation. Preserving principles of data sensitivity, data protection, and confidentiality requirements were met.

## Notes

### Clinical Protocols

https://bmjopen.bmj.com/content/bmjopen/8/10/e025896.full.pdf

### Author Declarations

The study had been approved by the Ethics Committee of the Medical Association Hamburg (study ID: PV5724).

### Summary of Updates

During the peer review process, we noticed an error in the order of the items of Table 2, Table 5 as well as Appendix 4 and Appendix 5. We corrected the order of the items (without altering any results).

## REFERENCES

1. Institute of Medicine. Crossing the Quality Chasm: A New Health System for the 21st Century. Washington, DC: The National Academies Press 2001.

2. Bodenheimer T, Sinsky C. From triple to quadruple aim: care of the patient requires care of the provider. The Annals of Family Medicine 2014;12(6):573–76.

3. Bertakis KD, Azari R. Patient-centered care is associated with decreased health care utilization. The Journal of the American Board of Family Medicine 2011;24(3):229–39.

4. Dwamena F, Holmes-Rovner M, Gaulden CM, et al. Interventions for providers to promote a patient-centred approach in clinical consultations. Cochrane Database of Systematic Reviews 2012(12) doi: 10.1002/14651858.CD003267.pub2

5. McMillan SS, Kendall E, Sav A, et al. Patient-centered approaches to health care: a systematic review of randomized controlled trials. Medical care research and review 2013;70(6):567–96.

6. Rathert C, Wyrwich MD, Boren SA. Patient-centered care and outcomes: a systematic review of the literature. Medical Care Research and Review 2013;70(4):351–79.

7. Bravo P, Härter M, McCaffery K, et al. 20 years after the start of international Shared Decision-Making activities: is it time to celebrate? Probably…: Elsevier, 2022:1–4.

8. Bundesministerium für Gesundheit. Gesundheitskompetenz und Patientenorientierung [Available from: https://www.bundesgesundheitsministerium.de/ministerium/ressortforschung/handlungsfelder/gesundheitskompetenz-und-patientenorientierung.html accessed 03.08.2023.

9. Dierks M-L, Seidel G, Schwartz FW, et al. Themenheft 32“ Bürger-und Patientenorientierung”. 2006

10. Bundesrat. Gesetz zur Verbesserung der Rechte von Patientinnen und Patienten [Available from: https://www.bundesaerztekammer.de/fileadmin/user_upload/_old-files/downloads/Patientenrechtegesetz_BGBl.pdf.

11. Brandstetter S, Curbach J, McCool M, et al. Patientenorientierung in der Versorgungsforschung. Das Gesundheitswesen 2014:200–05.

12. Arbeitskreis „Versorgungsforschung“ beim Wissenschaftlichen Beirat der Bundesärztekammer. Definition und Abgrenzung der Versorgungsforschung [Available from: https://www.bundesaerztekammer.de/fileadmin/user_upload/_old-files/downloads/pdf-Ordner/Versorgungsforschung/Definition.pdf.

13. Härter M, Dirmaier J, Scholl I, et al. The long way of implementing patient-centered care and shared decision making in Germany. *Zeitschrift für Evidenz*, Fortbildung und Qualität im Gesundheitswesen 2017;123:46–51.

14. Schmale-Grede R, Faubel U. Der Patient im Mittelpunkt der Versorgungsforschung. Zeitschrift für Rheumatologie 2020;79(10)

15. Wissenschaftsrat. Empfehlungen zur Weiterentwicklung des Medizinstudiums in Deutschland auf Grundlage einer Bestandsaufnahme der humanmedizinischen Modellstudiengänge Dresden2014 [Available from: https://www.wissenschaftsrat.de/download/archiv/4017-14.pdf?blob=publicationFile&v=1 accessed 26.08.2022.

16. Scholl I, Zill JM, Härter M, et al. An integrative model of patient-centeredness–a systematic review and concept analysis. PloS one 2014;9(9):e107828.

17. Zill JM, Scholl I, Härter M, et al. Which dimensions of patient-centeredness matter?-Results of a web-based expert delphi survey. PloS one 2015;10(11):e0141978.

18. Zeh S, Christalle E, Hahlweg P, et al. Assessing the relevance and implementation of patient-centredness from the patients’ perspective in Germany: results of a Delphi study. BMJ open 2019;9(12):e031741.

19. Epstein RM, Street RL. The values and value of patient-centered care: Annals Family Med, 2011:100–03.

20. Browne K, Roseman D, Shaller D, et al. Measuring patient experience as a strategy for improving primary care. Health affairs 2010;29(5):921–25.

21. Beattie M, Murphy DJ, Atherton I, et al. Instruments to measure patient experience of healthcare quality in hospitals: a systematic review. Systematic reviews 2015;4(1):1–21.

22. Coulter A. Can patients assess the quality of health care?: British Medical Journal Publishing Group, 2006:1-2.

23. Doyle C, Lennox L, Bell D. A systematic review of evidence on the links between patient experience and clinical safety and effectiveness. BMJ open 2013;3(1):e001570.

24. Anhang Price R, Elliott MN, Zaslavsky AM, et al. Examining the role of patient experience surveys in measuring health care quality. Medical Care Research and Review 2014;71(5):522–54.

25. Bull C, Byrnes J, Hettiarachchi R, et al. A systematic review of the validity and reliability of patient-reported experience measures. Health services research 2019;54(5):1023–35.

26. Mihaljevic AL, Doerr-Harim C, Kalkum E, et al. Measuring patient centeredness with German language Patient-Reported Experience Measures (PREM)–A systematic review and qualitative analysis according to COSMIN. Plos one 2022;17(11):e0264045.

27. Christalle E, Zeh S, Hahlweg P, et al. Development and content validity of the Experienced Patient-Centeredness Questionnaire (EPAT)—A best practice example for generating patient-reported measures from qualitative data. Health Expectations 2022

28. Christalle E, Zeh S, Hahlweg P, et al. Assessment of patient centredness through patient-reported experience measures (ASPIRED): protocol of a mixed-methods study. BMJ open 2018;8(10):e025896.

29. Gagnier JJ, Lai J, Mokkink LB, et al. COSMIN reporting guideline for studies on measurement properties of patient-reported outcome measures. Quality of Life Research 2021;30(8):2197–218.

30. Harkness J, Pennell BE, Schoua-Glusberg A. Survey questionnaire translation and assessment. Methods for testing and evaluating survey questionnaires 2004:453–73.

31. Wirtz MA, Morfeld M, Glaesmer H, et al. Konfirmatorische Prüfung der Skalenstruktur des SF-12 Version 2.0 in einer deutschen bevölkerungs-repräsentativen Stichprobe. Diagnostica 2018;64(2):84–96.

32. Kriz D, Nübling R, Steffanowski A, et al. Patientenzufriedenheit in der stationären Rehabilitation: Psychometrische Reanalyse des ZUF-8 auf der Basis multizentrischer Stichproben verschiedener Indikation. Zeitschrift für medizinische Psychologie 2008;17(2-3):67–79.

33. Röthlin F, Pelikan JM, Ganahl K. Die Gesundheitskompetenz der 15-jährigen Jugendlichen in Österreich. Abschlussbericht der österreichischen Gesundheitskompetenz Jugendstudie im Auftrag des Hauptverbands der österreichischen Sozialversicherungsträger (HVSV) 2013

34. Döring N, Bortz J. Forschungsmethoden und Evaluation. Wiesbaden: Springerverlag 2006

35. Boateng GO, Neilands TB, Frongillo EA, et al. Best practices for developing and validating scales for health, social, and behavioral research: a primer. Frontiers in public health 2018;6:149.

36. Rosseel Y. Lavaan estimators [Available from: https://lavaan.ugent.be/tutorial/est.html accessed 28.08.2022.

37. Irwing P, Booth T, Hughes DJ. The Wiley handbook of psychometric testing: A multidisciplinary reference on survey, scale and test development: John Wiley & Sons 2018.

38. Cochran WG. The χ2 test of goodness of fit. The Annals of mathematical statistics 1952:315–45.

39. Rodriguez A, Reise SP, Haviland MG. Evaluating bifactor models: Calculating and interpreting statistical indices. Psychological methods 2016;21(2):137.

40. Oltedal S, Garratt A, Bjertnæs Ø, et al. The NORPEQ patient experiences questionnaire: data quality, internal consistency and validity following a Norwegian inpatient survey. Scandinavian Journal of Public Health 2007;35(5):540–47.

41. Enders CK, Bandalos DL. The relative performance of full information maximum likelihood estimation for missing data in structural equation models. Structural equation modeling 2001;8(3):430–57.

42. Jordan S, Hoebel J. Health literacy of adults in Germany: Findings from the German Health Update (GEDA) study. Bundesgesundheitsblatt-Gesundheitsforschung-Gesundheitsschutz 2015;58:942–50.

43. Diamantopoulos A, Sarstedt M, Fuchs C, et al. Guidelines for choosing between multi-item and single-item scales for construct measurement: a predictive validity perspective. Journal of the Academy of Marketing Science 2012;40(3):434–49.

44. Chen FF, Hayes A, Carver CS, et al. Modeling general and specific variance in multifaceted constructs: A comparison of the bifactor model to other approaches. Journal of personality 2012;80(1):219–51.

45. Bull C, Callander EJ. Current PROM and PREM use in health system performance measurement: still a way to go. Patient Experience Journal 2022;9(1):12–18.

46. Reeves R, West E, Barron D. Facilitated patient experience feedback can improve nursing care: a pilot study for a phase III cluster randomised controlled trial. BMC Health Serv Res 2013;13:259. doi: 10.1186/1472-6963-13-259 [published Online First: 20130704]

47. Hibbard JH, Stockard J, Tusler M. Hospital performance reports: impact on quality, market share, and reputation. Health affairs 2005;24(4):1150–60.

48. Coulter A, Locock L, Ziebland S, et al. Collecting data on patient experience is not enough: they must be used to improve care. BMJ 2014;348:g2225. doi: 10.1136/bmj.g2225 [published Online First: 20140326]

49. Murante AM, Vainieri M, Rojas D, et al. Does feedback influence patient - professional communication? Empirical evidence from Italy. Health Policy 2014;116(2-3):273–80. doi: 10.1016/j.healthpol.2014.02.001 [published Online First: 20140224]

50. Eignor DR. The standards for educational and psychological testing. 2013

51. Zill JM, Lindig A, Reck LM, et al. Protocol: Assessment of person-centeredness in healthcare and social support services for women with unintended pregnancy (CarePreg): protocol for a mixed-method study. BMJ Open 2022;12(9)

