## Appendix 2 Confirmatory factor analyses for "Through the patients’ eyes - Psychometric evaluation of the 64-item version of the Experienced Patient-Centeredness Questionnaire (EPAT-64)"

**Appendix 2: Models tested in confirmatory factor analysis**

**Article:** Through the patients' eyes - Psychometric evaluation of the Experienced Patient-Centeredness Questionnaire (EPAT-64)

**Authors:** Eva Christalle, Stefan Zeh, Hannah Führes, Alica Schellhorn, Pola Hahlweg, Jördis Zill, Martin Härter, Carsten Bokemeyer, Jürgen Gallinat, Christoffer Gebhardt, Christina Magnussen, Volkmar Müller, Katharina Schmalstieg-Bahr, André Strahl, Levente Kriston, Isabelle Scholl

We developed items for each of the 16 dimensions of the integrative model of patient-centeredness.[1-3] Hence, we had a clear hypothesis which item should load on which dimension. We assumed that the 16 dimensions of PC are interrelated. Further, we hypothesized that there might be a general factor "patient-centeredness".

Based on those assumptions we tested five different models. Below you find a description of each model, including a sketch of the path model for two example dimensions (patient information and shared decision making). The real models included 16 specific dimensions.

**Model 1 – Unidimensional model**

- All items load on a single general factor.

*We tested this model to test whether the fit increases when we consider the 16 dimensions given in the integrative model of patient-centeredness.*

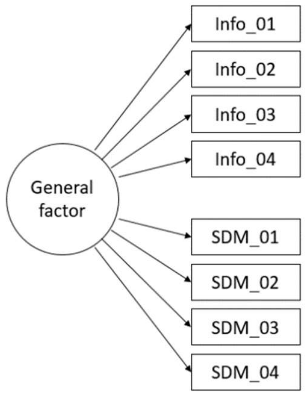


**Model 2 – Correlated first-order dimension model**

- All items load on their respective dimension.
- The dimensions correlate freely.

*This model is a direct translation of the integrative model of patient-centeredness. The model makes no assumptions about the interrelations of the dimensions or about the existence of a general factor. Comparison with the next models allows us to investigate whether the fit increases when we introduce a general factor.*

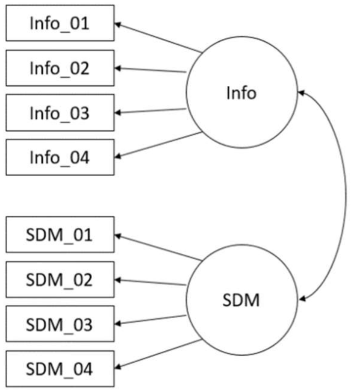

**Model 3 – Hierarchical model**

- All items load on their respective dimension.
- All dimensions load on a general factor.
- The dimensions correlate freely.

*In this model all dimensions are associated with the general factor. There is no direct association between the general factor and the items. The effect of the general factor on the items is modelled indirectly through the dimensions.*

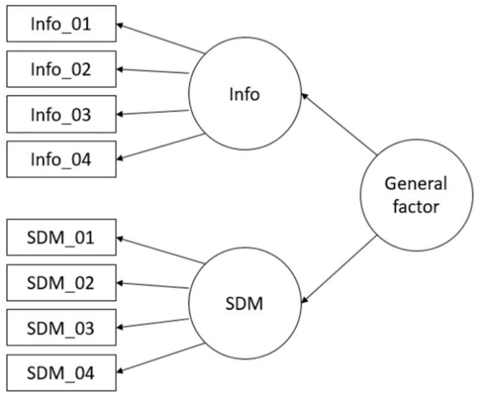


**Model 4 – Bifactor model with uncorrelated latent variables**

- All items load on their respective dimension.
- All items load on a general factor.
- Correlations of all dimensions and of dimensions and the general factor are restricted to 0.

*In contrast to a hierarchical model, bifactor models allows to model the role of the dimensions independently of the general factor.*[4] *This is the canonical form where the dimensions are not interrelated directly. Hence, all common variance between the dimensions are modelled solely by the influence of the general factor.*

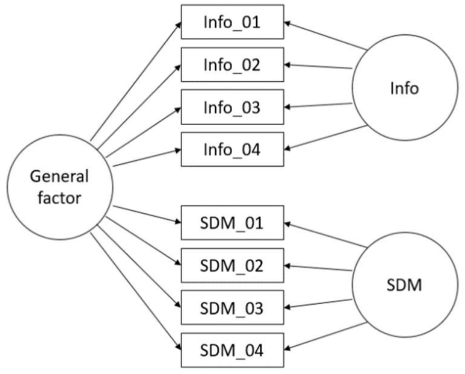


**Model 5 – Bifactor model with correlated dimension-specific latent variables**

- All items load on their respective dimension.
- All items load on a general factor.
- The dimensions correlate freely.
- Correlations of each dimension with the general factor are restricted to 0.

*In this bifactor model we again modelled the dimensions role independently of the general factor. Yet, we also postulated that there are direct relationships between the dimensions that are not accounted for by the general factor.*

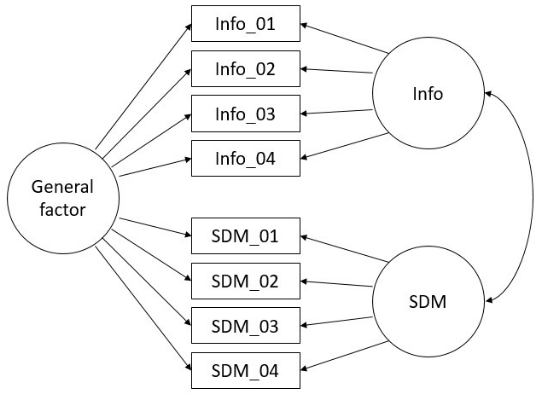


All models were estimated using the robust maximum likelihood estimator and full information maximum likelihood to deal with missing values.

References

1. Scholl I, Zill JM, Härter M, et al. An integrative model of patient-centeredness–a systematic review and concept analysis. *PloS one* 2014;9(9):e107828.
2. Zeh S, Christalle E, Hahlweg P, et al. Assessing the relevance and implementation of patient-centredness from the patients' perspective in Germany: results of a Delphi study. *BMJ open* 2019;9(12):e031741.
3. Christalle E, Zeh S, Hahlweg P, et al. Development and content validity of the Experienced Patient-Centeredness Questionnaire (EPAT)—A best practice example for generating patient-reported measures from qualitative data. *Health Expectations* 2022
4. Chen FF, West SG, Sousa KH. A comparison of bifactor and second-order models of quality of life. *Multivariate behavioral research* 2006;41(2):189-225.
