## Appendix 3 Subgroup sample characteristics for "Through the patients’ eyes - Psychometric evaluation of the 64-item version of the Experienced Patient-Centeredness Questionnaire (EPAT-64)"

**Appendix 3: Sample characteristics per disease group**

**Article:** Through the patients' eyes - Psychometric evaluation of the Experienced Patient-Centeredness Questionnaire (EPAT-64)

**Authors:** Eva Christalle, Stefan Zeh, Hannah Führes, Alica Schellhorn, Pola Hahlweg, Jördis Zill, Martin Härter, Carsten Bokemeyer, Jürgen Gallinat, Christoffer Gebhardt, Christina Magnussen, Volkmar Müller, Katharina Schmalstieg-Bahr, André Strahl, Levente Kriston, Isabelle Scholl

Table 1: Sample sizes per subgroup

| Disease group | Outpatients | | | Inpatients | | |
|---|---|---|---|---|---|---|
| | Paper | Online | Total | Paper | Online | Total |
| Cardiovascular | 269 | 8 | 277 | 260 | 26 | 286 |
| Cancer | 282 | 14 | 296 | 318 | 33 | 351 |
| Musculoskeletal | 116 | 101 | 218 | 29 | 63 | 92 |
| Mental | 192 | 80 | 272 | 77 | 125 | 202 |
| Total | 889 | 203 | 1092 | 684 | 247 | 931 |

Table 2: Sample characteristics per subgroup - outpatient sample

| Characteristics | Total | Cardiovascular diseases | Cancer | Musculoskeletal diseases | Mental disorders |
|---|---|---|---|---|---|
| **Sample size** | n = 1092 | n = 277 | n = 296 | n = 218 | n = 272 |
| **Age** (in years) | M = 53.1 (SD = 17.5) | M = 60.1 (SD = 17.8) | M = 59.2 (SD = 15.0) | M = 50.4 (SD = 15.7) | M = 42.6 (SD = 14.8) |
| **Years since diagnosis** | M = 11.4 (SD = 12.0) | M = 14.8 (SD = 13.5) | M = 5.8 (SD = 6.5) | M = 13.5 (SD = 13.9) | M = 12.6 (SD = 11.7) |
| **Years as patient in this outpatient clinic** | M = 4.53 (SD = 6.5) | M = 6.5 (SD = 9.4) | M = 2.7 (SD = 4.6) | M = 5.1 (SD = 6.5) | M = 3.6 (SD = 4.3) |
| **Health literacy**[a] | M = 50.7 (SD = 7.7) | M = 56.9 (SD = 7.5) | M = 50.8 (SD = 7.6) | M = 51.5 (SD = 7.3) | M = 50.0 (SD = 7.9) |
| **Satisfaction**[b] | M = 27.5 (SD = 4.6) | M = 28.1 (SD = 3.9) | M = 28.3 (SD = 3.8) | M = 26.3 (SD = 5.6) | M = 26.9 (SD = 4.8) |
| **Health status**[c] | M = 3.4 (SD = 2.8) | M = 3.1 (SD = 0.9) | M = 3.1 (SD = 0.8) | M = 3.4 (SD = 0.9) | M = 3.8 (SD = 5.3) |
| **Comorbidity** (Do you have any further diseases?) | n = 551 (55.0%) | n = 135 (55.3%) | n = 116 (46.0%) | n = 154 (75.1%) | n = 125 (48.8%) |
| **Gender** | | | | | |
| Female | n = 646 (60.3%) | n = 127 (46.4%) | n = 135 (50.9%) | n = 161 (76.3%) | n = 189 (69.7%) |
| Male | n = 420 (39.2%) | n = 147 (53.6%) | n = 129 (48.7%) | n = 48 (22.7%) | n = 80 (29.5%) |
| Diverse | n = 5 (0.5%) | n = 0 (0%) | n = 1 (0.4%) | n = 2 (0.9%) | n = 2 (0.7%) |
| **Marital status** | | | | | |
| Unmarried/unpartnered | n = 343 (32.3%) | n = 57 (20,8%) | n = 50 (19,0%) | n = 65 (31,0%) | n = 153 (57,7%) |
| Married/partnered | n = 552 (51.9%) | n = 169 (61,7%) | n = 173 (65,8%) | n = 117 (55,7%) | n = 71 (26,8%) |
| Divorced | n = 107 (10.1%) | n = 25 (9,1%) | n = 22 (8,4%) | n = 20 (9,5%) | n = 34 (12,8%) |
| Widowed | n = 61 (5.7%) | n = 23 (8,4%) | n = 18 (6,8%) | n = 8 (3,8%) | n = 7 (2,6%) |
| **Formal education** | | | | | |
| Low[d] | n = 10 (0.9%) | n = 33 (12.0%) | n = 46 (17.6%) | n = 16 (7.6%) | n = 21 (7.8%) |
| Intermediate[e] | n = 404 (37.8%) | n = 89 (32.4) | n = 67 (25.6%) | n = 58 (27.5%) | n = 61 (22.6%) |
| High[f] | n = 267 (25.0%) | n = 56 (20.4%) | n = 57 (21.8%) | n = 50 (23.7%) | n = 92 (34.0%) |
| Very high[g] | n = 376 (35.2%) | n = 93 (33.8%) | n = 90 (34.3%) | n = 84 (39.8%) | n = 93 (34.3%) |
| **Occupational status*** | | | | | |
| Employed | n = 447 (41.9%) | n = 96 (35.3%) | n = 110 (41.7%) | n = 110 (52.1%) | n = 106 (39.6%) |
| Unemployed | n = 66 (6.2%) | n = 5 (1.8%) | n = 14 (5.3%) | n = 7 (3.3%) | n = 39 (14.6%) |

| | | | | | |
|---|---|---|---|---|---|
| Student/trainee | n = 91 (8.5%) | n = 15 (5.5%) | n = 7 (2.7%) | n = 19 (9.0%) | n = 45 (16.5%) |
| Parental leave | n = 48 (4.5%) | n = 2 (0.7%) | n = 1 (0.4%) | n = 4 (1.8%) | n = 2 (0.7%) |
| Retired | n = 394 (37.0%) | n = 130 (47.8%) | n = 115 (43.6%) | n = 69 (32.7%) | n = 60 (22.4%) |
| **Health insurance*** | | | | | |
| Statutory | n = 910 (83.7%) | n = 211 (76.2%) | n = 216 (79.1%) | n = 190 (89.2%) | n = 246 (90.1%) |
| Private | n = 164 (15.1%) | n = 73 (26.4%) | n = 42 (15.4%) | n = 21 (9.9%) | n = 24 (8.8%) |
| **Migration background** (Were you or your parents born in another country than Germany?) | n = 167 (15.7%) | n = 24 (8.8%) | n = 37 (14.0%) | n = 43 (19.8%) | n = 54 (20.1%) |
| **Reason for visit in outpatient clinic*** | | | | | |
| Acute symptoms/emergency | n = 219 (23.4%) | n = 20 (13.9%) | n = 22 (8.4%) | n = 71 (33.6%) | n = 93 (34.6%) |
| Treatment planning | n = 246 (26.3%) | n = 33 (22.9%) | n = 80 (30.4%) | n = 51 (24.2%) | n = 72 (26.8%) |
| First appointment | n = 196 (20.9%) | n = 18 (12.5%) | n = 66 (25.1%) | n = 29 (13.4%) | n = 73 (27.1) |
| Carrying out treatment | n = 373 (39.8%) | n = 30 (20.8%) | n = 141 (53.6%) | n = 85 (40.3%) | n = 101 (37.5%) |
| Diagnostics/examination | n = 284 (30.3%) | n = 64 (44.4%) | n = 94 (35.7%) | n = 59 (28.0%) | n = 50 18.6%) |
| Progress discussion during treatment | n = 241 (25.7%) | n = 15 (10.4%) | n = 96 (36.5%) | n = 60 (28.4%) | n = 58 (21.6%) |
| Discussion of examination results | n = 209 (22.3%) | n = 33 (22.9%) | n = 100 (38.0%) | n = 40 (19.0%) | n = 19 (7.1%) |
| Follow-up | n = 301 (32.1%) | n = 79 (54.9%) | n = 92 (35%) | n = 65 (30.8%) | n = 48 (17.8%) |

**Note**: * multiple answers possible, M = Mean, SD = Standard Deviation, [a] Health literacy measured by HLS-EU-Q16(1), range 0-64, high value=high health literacy, [b] Treatment satisfaction measured by ZUF-8(2), range 8-32, high value=high satisfaction, [b] General health status measured by first item of SF-12(3), range 0-5, high value=low health status, [d] low = no formal degree or graduation after less than 10 years at school; [e] intermediate = graduation after 10 years at school; [f] high = graduation after more than 10 years at school; [g] very high = college or university degree

Table 3: Sample characteristics per subgroup - inpatient sample

| Characteristics | Total | Cardiovascular diseases | Cancer | Musculoskeletal diseases | Mental disorders |
|---|---|---|---|---|---|
| **Sample size** | n = 921 | n = 286 | n = 351 | n = 92 | n = 202 |
| **Age** (in years) | M = 56.0 (SD = 17.6) | M = 64.5 (SD = 14.0) | M = 60.9 (SD = 13.5) | M = 52.5 (SD = 16.4) | M = 37.0 (SD = 14.2) |
| **Years since diagnosis** | M = 8.5 (SD = 10.0) | M = 12.0 (SD = 11.3) | M = 3.7 (SD = 5.6) | M = 12.0 (SD = 12.4) | M = 9.9 (SD = 9.4) |
| **Length of stay** (in days) | M = 18.1 (SD = 30.6) | M = 7.0 (SD = 5.2) | M = 10.3 (SD = 11.4) | M = 16.9 (SD = 59.2) | M = 47.3 (SD = 37.2) |
| **Health literacy**[a] | M = 50.6 (SD = 7.7) | M = 46.5 (SD = 9.0) | M = 51.8 (SD = 7.4) | M = 50.2 (SD = 8.5) | M = 49.5 (SD = 7.3) |
| **Satisfaction**[b] | M = 28.0 (SD = 4.9) | M = 28.8 (SD = 4.0) | M = 29.7 (SD = 3.3) | M = 26.1 (SD = 5.6) | M = 24.7 (SD = 6.1) |
| **Health status**[c] | M = 3.4 (SD = 4.1) | M = 3.1 (SD = 0.8) | M = 3.1 (SD = 0.8) | M = 4.3 (SD = 9.0) | M = 4.0 (SD = 6.1) |
| **Comorbidity** (Do you have any further diseases?) | n = 467 (54.3%) | n = 130 (50.8%) | n = 159 (50.6%) | n = 62 (68.9%) | n = 107 (56.3%) |
| **Gender** | | | | | |
| Female | n = 384 (42.3%) | n = 105 (37.6%) | n = 93 (27.6%) | n = 57 (62.6%) | n = 124 (65.3%) |
| Male | n = 517 (57.0%) | n = 174 (62.4%) | n = 244 (72.4%) | n = 32 (35.2%) | n = 62 (32.6%) |
| Diverse | n = 6 (0.7%) | n = 0 (0%) | n = 0 (0%) | n = 2 (2.2%) | n = 4 (2.1%) |
| **Marital status** | | | | | |
| Unmarried/unpartnered | n = 237 (26.5%) | n = 27 (9.8%) | n = 49 (14.6%) | n = 27 (30.7%) | n = 132 (70.6%) |
| Married/partnered | n = 524 (58.5%) | n = 187 (68.0%) | n = 239 (71.3%) | n = 53 (60.2%) | n = 42 (22.5%) |
| Divorced | n = 80 (8.9%) | n = 30 (10.9%) | n = 27 (8.1%) | n = 7 (8.0%) | n = 12 (6.4%) |
| Widowed | n = 54 (6%) | n = 31 (11.3%) | n = 20 (6.0%) | n = 1 (1.1%) | n = 1 (0.5%) |
| **Formal education** | | | | | |
| Low[d] | n = 20 (2.2%) | n = 68 (24,5%) | n = 57 (17,0%) | n = 35 (39,3%) | n = 23 (11,9%) |
| Intermediate[e] | n = 383 (42.3%) | n = 82 (29,5%) | n = 75 (22,4%) | n = 6 (6,7%) | n = 57 (29,5%) |
| High[f] | n = 204 (22.6%) | n = 46 (16,5%) | n = 64 (19,1%) | n = 47 (52,8%) | n = 73 (37,8%) |
| Very high[g] | n = 279 (30.9%) | n = 75 (27,0%) | n = 131 (39,1%) | n = 1 (1,1%) | n = 38 (19,7%) |
| **Occupational status*** | | | | | |
| Employed | n = 357 (39.1%) | n = 98 (34.9%) | n = 143 (42.6%) | n = 38 (41.8%) | n = 76 (38.8%) |
| Unemployed | n = 64 (7.0%) | n = 11 (3.9%) | n = 8 (2.4%) | n = 4 (4.3%) | n = 41 (20.9%) |

| | | | | | |
|---|---|---|---|---|---|
| Student/trainee | n = 50 (5.5%) | n = 3 (1.1%) | n = 7 (2.1%) | n = 7 (7.7%) | n = 33 (16.8%) |
| Parental leave | n = 7 (0.8%) | n = 0 (0%) | n = 2 (0.6%) | n = 3 (3.3%) | n = 2 (1%) |
| Retired | n = 392 (42.9%) | n = 152 (54.1%) | n = 166 (49.4%) | n = 35 (38.5%) | n = 31 (15.8%) |
| **Health insurance*** | | | | | |
| Statutory | n = 721 (78.1%) | n = 223 (78.2%) | n = 226 (66.3%) | n = 78 (85.7%) | n = 184 (93.4%) |
| Private | n = 214 (23.2%) | n = 65 (22.8%) | n = 121 (35.6%) | n = 14 (15.2%) | n = 14 (7.1%) |
| **Migration background** (Were you or your parents born in another country than Germany?) | n = 150 (16.5%) | n = 31 (11.1%) | n = 50 (14.7%) | n = 13 (14.1%) | n = 55 (28.5%) |

**Note:** * multiple answers possible, M = Mean, SD = Standard Deviation, [a] Health literacy measured by HLS-EU-Q16(1), range 0-64, high value=high health literacy, [b] Treatment satisfaction measured by ZUF-8(2), range 8-32, high value=high satisfaction, [b] General health status measured by first item of SF-12(3), range 0-5, high value=low health status, [d] low = no formal degree or graduation after less than 10 years at school; [e] intermediate = graduation after 10 years at school; [f] high = graduation after more than 10 years at school; [g] very high = college or university degree

### References


1. Jordan S, Hoebel J. Health literacy of adults in Germany: Findings from the German Health Update (GEDA) study. Bundesgesundheitsblatt-Gesundheitsforschung-Gesundheitsschutz. 2015;58:942-50.
2. Kriz D, Nübling R, Steffanowski A, Wittmann WW, Schmidt J. Patientenzufriedenheit in der stationären Rehabilitation: Psychometrische Reanalyse des ZUF-8 auf der Basis multizentrischer Stichproben verschiedener Indikation. Zeitschrift für medizinische Psychologie. 2008;17(2-3):67-79.
3. Wirtz MA, Morfeld M, Glaesmer H, Brähler E. Konfirmatorische Prüfung der Skalenstruktur des SF-12 Version 2.0 in einer deutschen bevölkerungs-repräsentativen Stichprobe. Diagnostica. 2018;64(2):84-96.
