## Appendix 4 Item characteristics per medical condition - outpatient sample for "Through the patients’ eyes - Psychometric evaluation of the 64-item version of the Experienced Patient-Centeredness Questionnaire (EPAT-64)"

**Appendix 4: Item characteristics per disease group - outpatient sample**

**Article:** Through the patients' eyes - Psychometric evaluation of the Experienced Patient-Centeredness Questionnaire (EPAT-64)

**Authors:** Eva Christalle, Stefan Zeh, Hannah Führes, Alica Schellhorn, Pola Hahlweg, Jördis Zill, Martin Härter, Carsten Bokemeyer, Jürgen Gallinat, Christoffer Gebhardt, Christina Magnussen, Volkmar Müller, Katharina Schmalstieg-Bahr, André Strahl, Levente Kriston, Isabelle Scholl

**Abbreviations:**

- Card = Cardiovascular diseases
- Mus = Muscoloskeletal diseases
- Ment = Mental disorders

**Dimension “Essential characteristics of the clinicians”**

| | | mean | | | | | standard deviation | | | | |
|---|---|---|---|---|---|---|---|---|---|---|---|
| | Item | **All** | Card | Cancer | Mus | Ment | **All** | Card | Cancer | Mus | Ment |
| Item 1 | The healthcare professionals were sensitive (for example they addressed my feelings, showed understanding, or empathized with my situation). | **5.1** | 5.1 | 5.1 | 4.8 | 5.1 | **1.2** | 1.2 | 1.2 | 1.5 | 1.2 |
| Item 2 | The healthcare professionals behaved respectfully and appreciatively. | **5.5** | 5.6 | 5.6 | 5.3 | 5.6 | **0.9** | 0.8 | 0.7 | 1.1 | 0.7 |
| Item 3 | The healthcare professionals were committed to finding a solution for my health concerns. | **5.2** | 5.3 | 5.3 | 5.0 | 5.3 | **1.1** | 1.0 | 1.0 | 1.3 | 1.0 |
| Item 4 | If I wanted to, difficult topics were discussed directly and openly by the healthcare professionals (for example, long-term effects of the illness, life expectancy, or sexuality). | **4.8** | 4.7 | 4.9 | 4.4 | 4.9 | **1.5** | 1.6 | 1.4 | 1.6 | 1.4 |

| | Item difficulty | | | | | 'does not concern me' | | | | | no reply | | | | | item total correlation | | | | |
|---|---|---|---|---|---|---|---|---|---|---|---|---|---|---|---|---|---|---|---|---|
| | **All** | Card | Cancer | Mus | Ment | **All** | Card | Cancer | Mus | Ment | **All** | Card | Cancer | Mus | Ment | **All** | Card | Cancer | Mus | Ment |
| Item 1 | **82.1** | 81.5 | 82.2 | 75.2 | 82.2 | **4.2%** | 8.3% | 4.4% | 2.8% | 4.4% | **1.1%** | 1.4% | 1.8% | 0.5% | 1.8% | **0.80** | 0.73 | 0.76 | 0.88 | 0.76 |
| Item 2 | **90.8** | 91.5 | 92.8 | 86.3 | 92.8 | **0.7%** | 1.1% | 0.7% | 0.0% | 0.7% | **0.7%** | 0.4% | 1.5% | 0.5% | 1.5% | **0.73** | 0.69 | 0.65 | 0.83 | 0.65 |
| Item 3 | **85.0** | 86.4 | 86.7 | 80.2 | 86.7 | **4.5%** | 10.1% | 5.1% | 1.4% | 5.1% | **0.8%** | 0.4% | 1.5% | 0.5% | 1.5% | **0.80** | 0.78 | 0.76 | 0.86 | 0.76 |
| Item 4 | **76.9** | 74.7 | 78.6 | 68.2 | 78.6 | **32.8%** | 42.2% | 27.8% | 36.4% | 27.8% | **1.1%** | 0.7% | 2.2% | 0.5% | 2.2% | **0.71** | 0.66 | 0.71 | 0.80 | 0.71 |

**Dimension "Clinician-patient relationship"**

| | | mean | | | | | standard deviation | | | | |
|---|---|---|---|---|---|---|---|---|---|---|---|
| | Item | **All** | Card | Cancer | Mus | Ment | **All** | Card | Cancer | Mus | Ment |
| Item 1 | I trusted my healthcare professionals. | **5.2** | 5.3 | 5.3 | 5.0 | 5.3 | **1.1** | 1.1 | 1.0 | 1.3 | 1.0 |
| Item 2 | I felt I could confide in my healthcare professionals (for example, on intimate or difficult topics). | **4.9** | 4.8 | 4.8 | 4.5 | 4.8 | **1.4** | 1.5 | 1.3 | 1.6 | 1.3 |
| Item 3 | The healthcare professionals knew about my medical history and my current health status. | **5.1** | 5.2 | 5.3 | 4.8 | 5.3 | **1.2** | 1.1 | 0.9 | 1.4 | 0.9 |
| Item 4 | Existing complaints were addressed again in follow-up meetings. | **5.0** | 4.8 | 5.1 | 4.7 | 5.1 | **1.3** | 1.5 | 1.2 | 1.5 | 1.2 |

| | Item difficulty | | | | | 'does not concern me' | | | | | no reply | | | | | item total correlation | | | | |
|---|---|---|---|---|---|---|---|---|---|---|---|---|---|---|---|---|---|---|---|---|
| | **All** | Card | Cancer | Mus | Ment | **All** | Card | Cancer | Mus | Ment | **All** | Card | Cancer | Mus | Ment | **All** | Card | Cancer | Mus | Ment |
| Item 1 | **84.8** | 86.0 | 86.1 | 79.2 | 86.1 | **1.8%** | 4.7% | 0.4% | 0.9% | 0.4% | **1.2%** | 1.1% | 1.5% | 0.9% | 1.5% | **0.77** | 0.70 | 0.78 | 0.82 | 0.78 |
| Item 2 | **77.8** | 76.0 | 77.0 | 70.7 | 77.0 | **12.8%** | 26.7% | 10.6% | 11.5% | 10.6% | **1.0%** | 0.7% | 1.5% | 0.5% | 1.5% | **0.75** | 0.71 | 0.74 | 0.79 | 0.74 |
| Item 3 | **82.2** | 83.0 | 86.2 | 77.0 | 86.2 | **3.8%** | 6.5% | 1.1% | 2.3% | 1.1% | **0.5%** | 0.0% | 1.1% | 0.5% | 1.1% | **0.57** | 0.56 | 0.55 | 0.65 | 0.55 |
| Item 4 | **79.2** | 75.2 | 81.7 | 74.0 | 81.7 | **23.0%** | 33.6% | 21.2% | 15.2% | 21.2% | **0.7%** | 0.4% | 1.5% | 0.5% | 1.5% | **0.68** | 0.59 | 0.63 | 0.77 | 0.63 |

**Dimension "Patient as a unique person"**

| | Item | mean | | | | | standard deviation | | | | |
|---|---|---|---|---|---|---|---|---|---|---|---|
| | | **All** | Card | Cancer | Mus | Ment | **All** | Card | Cancer | Mus | Ment |
| Item 1 | My wishes, needs and expectations were asked and taken into account in the treatment. | **4.8** | 4.7 | 4.7 | 4.5 | 4.7 | **1.4** | 1.4 | 1.4 | 1.6 | 1.4 |
| Item 2 | My healthcare professionals addressed me personally and did not treat me as just one of many patients. | **5.1** | 5.1 | 5.1 | 4.8 | 5.1 | **1.2** | 1.1 | 1.2 | 1.4 | 1.2 |
| Item 3 | My personal health goals were asked and taken into account. | **4.1** | 3.7 | 3.8 | 3.9 | 3.8 | **1.8** | 1.9 | 1.7 | 1.8 | 1.7 |
| Item 4 | It was asked and taken into account what opportunities and skills I can provide to support my health. | **3.7** | 3.2 | 3.4 | 3.6 | 3.4 | **1.8** | 1.8 | 1.8 | 1.9 | 1.8 |

| | Item difficulty | | | | | 'does not concern me' | | | | | no reply | | | | | item total correlation | | | | |
|---|---|---|---|---|---|---|---|---|---|---|---|---|---|---|---|---|---|---|---|---|
| | **All** | Card | Cancer | Mus | Ment | **All** | Card | Cancer | Mus | Ment | **All** | Card | Cancer | Mus | Ment | **All** | Card | Cancer | Mus | Ment |
| Item 1 | **75.1** | 74.5 | 73.3 | 69.5 | 73.3 | **8.3%** | 14.1% | 10.6% | 3.7% | 10.6% | **0.8%** | 1.1% | 0.7% | 1.4% | 0.7% | **0.77** | 0.71 | 0.74 | 0.88 | 0.74 |
| Item 2 | **82.1** | 82.6 | 82.0 | 76.8 | 82.0 | **2.0%** | 4.0% | 1.8% | 0.5% | 1.8% | **0.5%** | 0.0% | 0.7% | 1.4% | 0.7% | **0.66** | 0.55 | 0.62 | 0.78 | 0.62 |
| Item 3 | **62.4** | 54.4 | 55.1 | 58.9 | 55.1 | **12.5%** | 23.5% | 13.9% | 7.4% | 13.9% | **0.7%** | 0.7% | 0.4% | 1.4% | 0.4% | **0.76** | 0.73 | 0.74 | 0.81 | 0.74 |
| Item 4 | **54.9** | 44.5 | 48.4 | 51.8 | 48.4 | **17.7%** | 31.8% | 17.6% | 12.4% | 17.6% | **0.7%** | 0.4% | 0.7% | 1.4% | 0.7% | **0.72** | 0.68 | 0.73 | 0.74 | 0.73 |

**Dimension “Biopsychosocial perspective”**

| | | mean | | | | | standard deviation | | | | |
|---|---|---|---|---|---|---|---|---|---|---|---|
| | Item | **All** | Card | Cancer | Mus | Ment | **All** | Card | Cancer | Mus | Ment |
| Item 1 | My entire personal life was taken into account during the treatment (for example, job, family and friends, partnership and sexuality, culture and religion, age, or financial circumstances). | **3.9** | 3.2 | 3.5 | 3.4 | 3.5 | **1.8** | 1.8 | 1.8 | 1.8 | 1.8 |
| Item 2 | I was asked how my condition affects my life. | **4.0** | 4.1 | 3.5 | 3.4 | 3.5 | **1.8** | 1.8 | 1.8 | 1.9 | 1.8 |
| Item 3 | My entire medical history was asked and taken into account. | **4.9** | 4.9 | 4.8 | 4.6 | 4.8 | **1.4** | 1.4 | 1.5 | 1.5 | 1.5 |
| Item 4 | I was informed about the interaction of physical, psychological, and social factors. | **3.4** | 2.6 | 3.3 | 2.9 | 3.3 | **1.9** | 1.8 | 1.8 | 1.9 | 1.8 |

| | Item difficulty | | | | | 'does not concern me' | | | | | no reply | | | | | item total correlation | | | | |
|---|---|---|---|---|---|---|---|---|---|---|---|---|---|---|---|---|---|---|---|---|
| | **All** | Card | Cancer | Mus | Ment | **All** | Card | Cancer | Mus | Ment | **All** | Card | Cancer | Mus | Ment | **All** | Card | Cancer | Mus | Ment |
| Item 1 | **57.0** | 43.9 | 49.4 | 48.6 | 49.4 | **16.1%** | 28.9% | 18.3% | 12.4% | 18.3% | **1.2%** | 1.8% | 1.8% | 0.5% | 1.8% | **0.77** | 0.72 | 0.80 | 0.78 | 0.80 |
| Item 2 | **60.9** | 61.6 | 50.4 | 48.9 | 50.4 | **10.1%** | 17.3% | 10.6% | 8.8% | 10.6% | **1.6%** | 1.8% | 1.5% | 0.9% | 1.5% | **0.74** | 0.64 | 0.74 | 0.73 | 0.74 |
| Item 3 | **77.1** | 78.2 | 76.0 | 71.3 | 76.0 | **7.9%** | 13.7% | 9.9% | 3.7% | 9.9% | **1.5%** | 1.8% | 2.2% | 0.5% | 2.2% | **0.60** | 0.56 | 0.56 | 0.59 | 0.56 |
| Item 4 | **47.6** | 32.9 | 45.8 | 37.3 | 45.8 | **20.2%** | 36.5% | 16.5% | 16.1% | 16.5% | **1.1%** | 1.4% | 1.8% | 0.5% | 1.8% | **0.73** | 0.68 | 0.74 | 0.72 | 0.74 |

**Dimension “Clinician-patient communication”**

| | | mean | | | | | standard deviation | | | | |
|---|---|---|---|---|---|---|---|---|---|---|---|
| | Item | **All** | Card | Cancer | Mus | Ment | **All** | Card | Cancer | Mus | Ment |
| Item 1 | I was given enough time to describe my concerns and my situation (for example, medical history or current symptoms). | **5.3** | 5.3 | 5.4 | 5.1 | 5.4 | **1.1** | 1.2 | 0.9 | 1.3 | 0.9 |
| Item 2 | The healthcare professionals used terms that were easy to understand. | **5.4** | 5.2 | 5.3 | 5.3 | 5.3 | **1.0** | 1.0 | 0.9 | 1.1 | 0.9 |
| Item 3 | The healthcare professionals looked at me and listened carefully during our conversation. | **5.5** | 5.4 | 5.5 | 5.2 | 5.5 | **0.9** | 1.0 | 0.8 | 1.2 | 0.8 |
| Item 4 | The healthcare professionals ensured that I understood correctly what was explained to me. | **5.0** | 5.1 | 5.1 | 4.5 | 5.1 | **1.3** | 1.2 | 1.1 | 1.6 | 1.1 |

| | Item difficulty | | | | | 'does not concern me' | | | | | no reply | | | | | item total correlation | | | | |
|---|---|---|---|---|---|---|---|---|---|---|---|---|---|---|---|---|---|---|---|---|
| | **All** | Card | Cancer | Mus | Ment | **All** | Card | Cancer | Mus | Ment | **All** | Card | Cancer | Mus | Ment | **All** | Card | Cancer | Mus | Ment |
| Item 1 | **86.3** | 85.9 | 87.9 | 81.1 | 87.9 | **2.6%** | 5.1% | 2.2% | 2.3% | 2.2% | **1.0%** | 2.5% | 1.1% | 0.5% | 1.1% | **0.73** | 0.74 | 0.67 | 0.81 | 0.67 |
| Item 2 | **87.4** | 84.8 | 85.6 | 86.3 | 85.6 | **0.8%** | 1.8% | 0.4% | 0.5% | 0.4% | **0.8%** | 1.1% | 0.7% | 0.9% | 0.7% | **0.58** | 0.64 | 0.58 | 0.60 | 0.58 |
| Item 3 | **89.9** | 88.7 | 90.7 | 83.6 | 90.7 | **0.4%** | 0.4% | 0.4% | 0.5% | 0.4% | **0.5%** | 0.7% | 0.7% | 0.5% | 0.7% | **0.77** | 0.79 | 0.73 | 0.81 | 0.73 |
| Item 4 | **79.6** | 81.2 | 81.9 | 70.1 | 81.9 | **5.2%** | 6.1% | 4.8% | 4.6% | 4.8% | **0.9%** | 1.8% | 1.1% | 0.9% | 1.1% | **0.67** | 0.73 | 0.67 | 0.65 | 0.67 |

**Dimension "Integration of medical and non-medical care"**

| | | mean | | | | | standard deviation | | | | |
|---|---|---|---|---|---|---|---|---|---|---|---|
| | Item | **All** | Card | Cancer | Mus | Ment | **All** | Card | Cancer | Mus | Ment |
| Item 1 | I was asked if I use or would like to use additional services (for example, support groups, counseling, health courses, complementary and alternative medicine, or spiritual support/pastoral care). | **3.1** | 2.3 | 3.0 | 2.6 | 3.0 | **2.0** | 1.9 | 1.9 | 1.8 | 1.9 |
| Item 2 | If I used or wanted to use additional services, it was accepted. | **4.5** | 3.4 | 4.4 | 4.5 | 4.4 | **1.8** | 2.1 | 1.9 | 1.7 | 1.9 |
| Item 3 | The healthcare professionals informed me about the advantages and disadvantages of additional services. | **3.1** | 2.5 | 2.9 | 2.9 | 2.9 | **1.9** | 1.9 | 1.9 | 2.0 | 1.9 |
| Item 4 | If necessary, I was given specific contacts where I could get information about additional offers. | **3.3** | 2.6 | 3.2 | 2.5 | 3.2 | **2.1** | 2.1 | 2.1 | 1.7 | 2.1 |

| | Item difficulty | | | | | 'does not concern me' | | | | | no reply | | | | | item total correlation | | | | |
|---|---|---|---|---|---|---|---|---|---|---|---|---|---|---|---|---|---|---|---|---|
| | **All** | Card | Cancer | Mus | Ment | **All** | Card | Cancer | Mus | Ment | **All** | Card | Cancer | Mus | Ment | **All** | Card | Cancer | Mus | Ment |
| Item 1 | **42.3** | 26.9 | 39.5 | 31.5 | 39.5 | **30.4%** | 54.5% | 23.8% | 29.0% | 23.8% | **1.1%** | 1.8% | 1.8% | 0.5% | 1.8% | **0.79** | 0.82 | 0.82 | 0.74 | 0.82 |
| Item 2 | **70.6** | 47.8 | 67.3 | 69.8 | 67.3 | **57.6%** | 73.6% | 62.3% | 55.3% | 62.3% | **1.7%** | 3.2% | 2.6% | 0.5% | 2.6% | **0.73** | 0.86 | 0.74 | 0.68 | 0.74 |
| Item 3 | **42.0** | 29.7 | 38.5 | 37.9 | 38.5 | **43.3%** | 63.2% | 39.9% | 42.9% | 39.9% | **1.7%** | 2.5% | 3.3% | 0.5% | 3.3% | **0.78** | 0.86 | 0.80 | 0.77 | 0.80 |
| Item 4 | **45.3** | 32.7 | 43.4 | 29.4 | 43.4 | **46.2%** | 64.6% | 44.3% | 48.4% | 44.3% | **1.9%** | 2.5% | 3.3% | 0.5% | 3.3% | **0.80** | 0.85 | 0.87 | 0.69 | 0.87 |

**Dimension "Teamwork and teambuilding"**

| | | mean | | | | | standard deviation | | | | |
|---|---|---|---|---|---|---|---|---|---|---|---|
| | Item | **All** | Card | Cancer | Mus | Ment | **All** | Card | Cancer | Mus | Ment |
| Item 1 | The processes within the team were well organized. | **5.1** | 5.0 | 5.2 | 5.2 | 5.2 | **1.0** | 1.1 | 1.0 | 0.9 | 1.0 |
| Item 2 | The entire outpatient clinic team was responsible and approachable for me. | **5.0** | 5.1 | 5.3 | 4.8 | 5.3 | **1.2** | 1.1 | 0.9 | 1.4 | 0.9 |
| Item 3 | The outpatient clinic team exchanged information about my current health status (for example, everyone was informed about test results). | **4.6** | 4.7 | 4.9 | 4.2 | 4.9 | **1.4** | 1.3 | 1.2 | 1.6 | 1.2 |
| Item 4 | Various healthcare professionals within the outpatient clinic team have given me contradictory information. | **5.1** | 5.2 | 5.1 | 5.0 | 5.1 | **1.4** | 1.4 | 1.4 | 1.4 | 1.4 |

| | Item difficulty | | | | | 'does not concern me' | | | | | no reply | | | | | item total correlation | | | | |
|---|---|---|---|---|---|---|---|---|---|---|---|---|---|---|---|---|---|---|---|---|
| | **All** | Card | Cancer | Mus | Ment | **All** | Card | Cancer | Mus | Ment | **All** | Card | Cancer | Mus | Ment | **All** | Card | Cancer | Mus | Ment |
| Item 1 | **82.8** | 80.8 | 83.9 | 83.6 | 83.9 | **7.1%** | 3.6% | 3.7% | 2.8% | 3.7% | **0.8%** | 1.8% | 0.7% | 0.5% | 0.7% | **0.53** | 0.55 | 0.51 | 0.57 | 0.51 |
| Item 2 | **80.3** | 81.4 | 85.5 | 75.7 | 85.5 | **12.4%** | 9.0% | 5.9% | 6.5% | 5.9% | **1.4%** | 2.5% | 1.5% | 0.5% | 1.5% | **0.59** | 0.60 | 0.64 | 0.51 | 0.64 |
| Item 3 | **72.0** | 73.9 | 77.2 | 63.3 | 77.2 | **22.2%** | 15.9% | 10.6% | 20.3% | 10.6% | **2.1%** | 2.9% | 2.9% | 1.4% | 2.9% | **0.54** | 0.52 | 0.51 | 0.53 | 0.51 |
| Item 4 | **81.7** | 83.5 | 82.3 | 80.7 | 82.3 | **26.1%** | 21.3% | 14.7% | 23.0% | 14.7% | **0.9%** | 1.4% | 1.1% | 0.5% | 1.1% | **0.28** | 0.21 | 0.27 | 0.36 | 0.27 |

**Dimension "Access to care"**

| | Item | mean | | | | | standard deviation | | | | |
|---|---|---|---|---|---|---|---|---|---|---|---|
| | | **All** | Card | Cancer | Mus | Ment | **All** | Card | Cancer | Mus | Ment |
| Item 1 | If I wanted to speak to a physician, they were easily accessible. | **4.7** | 4.6 | 5.0 | 4.3 | 5.0 | **1.4** | 1.4 | 1.0 | 1.6 | 1.0 |
| Item 2 | I received an appointment in time. | **5.1** | 5.2 | 5.6 | 4.7 | 5.6 | **1.2** | 1.2 | 0.7 | 1.5 | 0.7 |
| Item 3 | I was able to easily schedule an appointment at the outpatient clinic (for example, via telephone, e-mail, or website). | **5.0** | 5.1 | 5.1 | 4.8 | 5.1 | **1.4** | 1.3 | 1.3 | 1.5 | 1.3 |
| Item 4 | The scheduled appointments at the outpatient clinic were conveniently timed for me (for example, compatible with work or school). | **5.1** | 5.1 | 5.4 | 5.0 | 5.4 | **1.3** | 1.3 | 1.0 | 1.4 | 1.0 |

| | Item difficulty | | | | | 'does not concern me' | | | | | no reply | | | | | item total correlation | | | | |
|---|---|---|---|---|---|---|---|---|---|---|---|---|---|---|---|---|---|---|---|---|
| | **All** | Card | Cancer | Mus | Ment | **All** | Card | Cancer | Mus | Ment | **All** | Card | Cancer | Mus | Ment | **All** | Card | Cancer | Mus | Ment |
| Item 1 | **73.1** | 72.6 | 80.4 | 66.0 | 80.4 | **23.4%** | 30.7% | 12.8% | 33.6% | 12.8% | **1.3%** | 1.4% | 2.2% | 0.5% | 2.2% | **0.54** | 0.55 | 0.40 | 0.48 | 0.40 |
| Item 2 | **82.5** | 83.9 | 91.8 | 73.4 | 91.8 | **2.1%** | 3.6% | 0.7% | 0.9% | 0.7% | **1.4%** | 2.5% | 1.5% | 0.9% | 1.5% | **0.62** | 0.55 | 0.53 | 0.67 | 0.53 |
| Item 3 | **79.5** | 81.7 | 82.3 | 75.1 | 82.3 | **9.2%** | 15.5% | 8.8% | 4.1% | 8.8% | **1.3%** | 1.8% | 2.2% | 0.5% | 2.2% | **0.57** | 0.59 | 0.39 | 0.60 | 0.39 |
| Item 4 | **82.7** | 81.2 | 88.1 | 79.1 | 88.1 | **8.6%** | 13.0% | 7.7% | 4.6% | 7.7% | **0.9%** | 1.1% | 1.5% | 0.9% | 1.5% | **0.50** | 0.52 | 0.37 | 0.56 | 0.37 |

**Dimension "Coordination and continuity of care"**

| | | mean | | | | | standard deviation | | | | |
|---|---|---|---|---|---|---|---|---|---|---|---|
| | Item | **All** | Card | Cancer | Mus | Ment | **All** | Card | Cancer | Mus | Ment |
| Item 1 | It was discussed with me whether follow-up appointments would be useful (for example, for aftercare or further treatment). | **5.1** | 5.2 | 5.2 | 4.8 | 5.2 | **1.4** | 1.3 | 1.2 | 1.6 | 1.2 |
| Item 2 | I was explained how long I will approximately have to wait and why. | **3.9** | 3.4 | 3.8 | 3.9 | 3.8 | **1.8** | 1.9 | 1.7 | 1.8 | 1.7 |
| Item 3 | The healthcare professionals took enough time for me. | **5.3** | 5.3 | 5.4 | 4.9 | 5.4 | **1.1** | 1.0 | 0.9 | 1.3 | 0.9 |
| Item 4 | Treatment steps were recorded in my treatment plan. | **3.5** | 3.8 | 4.3 | 2.8 | 4.3 | **2.0** | 1.9 | 1.8 | 1.9 | 1.8 |

| | Item difficulty | | | | | 'does not concern me' | | | | | no reply | | | | | item total correlation | | | | |
|---|---|---|---|---|---|---|---|---|---|---|---|---|---|---|---|---|---|---|---|---|
| | **All** | Card | Cancer | Mus | Ment | **All** | Card | Cancer | Mus | Ment | **All** | Card | Cancer | Mus | Ment | **All** | Card | Cancer | Mus | Ment |
| Item 1 | **81.7** | 83.5 | 84.6 | 76.4 | 84.6 | **13.1%** | 16.2% | 12.8% | 11.1% | 12.8% | **1.4%** | 0.7% | 3.3% | 0.0% | 3.3% | **0.43** | 0.37 | 0.46 | 0.42 | 0.46 |
| Item 2 | **57.7** | 47.8 | 55.0 | 58.7 | 55.0 | **11.0%** | 7.6% | 9.2% | 13.8% | 9.2% | **0.9%** | 0.4% | 2.2% | 0.0% | 2.2% | **0.35** | 0.36 | 0.40 | 0.40 | 0.40 |
| Item 3 | **85.1** | 85.8 | 87.5 | 78.6 | 87.5 | **0.5%** | 0.7% | 0.7% | 0.5% | 0.7% | **0.9%** | 0.4% | 2.2% | 0.5% | 2.2% | **0.48** | 0.49 | 0.43 | 0.51 | 0.43 |
| Item 4 | **50.1** | 55.4 | 65.5 | 35.7 | 65.5 | **27.2%** | 34.7% | 19.8% | 24.9% | 19.8% | **1.5%** | 2.9% | 1.8% | 0.5% | 1.8% | **0.43** | 0.46 | 0.52 | 0.41 | 0.52 |

**Dimension “Patient safety”**

| | | mean | | | | | standard deviation | | | | |
|---|---|---|---|---|---|---|---|---|---|---|---|
| | Item | **All** | Card | Cancer | Mus | Ment | **All** | Card | Cancer | Mus | Ment |
| Item 1 | I was encouraged to speak up if I noticed inconsistencies in my treatment. | **3.9** | 3.7 | 4.1 | 3.5 | 4.1 | **1.8** | 1.9 | 1.8 | 1.9 | 1.8 |
| Item 2 | I was examined thoroughly and carefully. | **5.1** | 5.4 | 5.2 | 4.9 | 5.2 | **1.2** | 1.0 | 1.2 | 1.4 | 1.2 |
| Item 3 | When I was prescribed new medication, I was asked what other medication I am taking and whether I have any intolerances. | **5.0** | 5.2 | 5.2 | 4.4 | 5.2 | **1.5** | 1.5 | 1.2 | 1.8 | 1.2 |
| Item 4 | I was informed about whom to contact if there was an inconsistency in my treatment or if I wanted to file a complaint. | **2.3** | 2.1 | 2.6 | 2.0 | 2.6 | **1.7** | 1.6 | 1.7 | 1.6 | 1.7 |

| | Item difficulty | | | | | 'does not concern me' | | | | | no reply | | | | | item total correlation | | | | |
|---|---|---|---|---|---|---|---|---|---|---|---|---|---|---|---|---|---|---|---|---|
| | **All** | Card | Cancer | Mus | Ment | **All** | Card | Cancer | Mus | Ment | **All** | Card | Cancer | Mus | Ment | **All** | Card | Cancer | Mus | Ment |
| Item 1 | **57.5** | 54.1 | 61.6 | 50.6 | 61.6 | **23.4%** | 31.0% | 18.3% | 18.4% | 18.3% | **0.8%** | 0.7% | 1.5% | 0.9% | 1.5% | **0.57** | 0.47 | 0.68 | 0.67 | 0.68 |
| Item 2 | **81.9** | 88.9 | 84.1 | 77.5 | 84.1 | **16.6%** | 5.8% | 10.3% | 9.2% | 10.3% | **2.7%** | 3.6% | 2.9% | 2.3% | 2.9% | **0.41** | 0.25 | 0.44 | 0.50 | 0.44 |
| Item 3 | **79.7** | 83.2 | 84.9 | 68.4 | 84.9 | **41.6%** | 51.3% | 29.3% | 40.1% | 29.3% | **1.0%** | 1.4% | 1.1% | 0.5% | 1.1% | **0.54** | 0.40 | 0.58 | 0.63 | 0.58 |
| Item 4 | **25.9** | 21.8 | 32.0 | 19.9 | 32.0 | **30.7%** | 39.0% | 24.2% | 27.2% | 24.2% | **0.9%** | 1.8% | 1.1% | 0.5% | 1.1% | **0.47** | 0.47 | 0.50 | 0.47 | 0.50 |

**Dimension "Patient information"**

| | Item | mean | | | | | standard deviation | | | | |
|---|---|---|---|---|---|---|---|---|---|---|---|
| | | **All** | Card | Cancer | Mus | Ment | **All** | Card | Cancer | Mus | Ment |
| Item 1 | I received information about my condition from my healthcare professionals (for example, causes, symptoms, effects or course). | **4.6** | 4.6 | 4.8 | 4.3 | 4.8 | **1.6** | 1.6 | 1.4 | 1.6 | 1.4 |
| Item 2 | I was asked what I already know about my condition. | **3.9** | 3.9 | 3.8 | 3.5 | 3.8 | **1.8** | 1.8 | 1.7 | 1.9 | 1.7 |
| Item 3 | The significance of my test results was explained to me. | **4.8** | 4.8 | 5.1 | 4.5 | 5.1 | **1.5** | 1.6 | 1.2 | 1.6 | 1.2 |
| Item 4 | I was asked what I would like to know about my condition. | **4.0** | 4.1 | 4.5 | 3.6 | 4.5 | **1.8** | 1.8 | 1.6 | 1.9 | 1.6 |

| | Item difficulty | | | | | 'does not concern me' | | | | | no reply | | | | | item total correlation | | | | |
|---|---|---|---|---|---|---|---|---|---|---|---|---|---|---|---|---|---|---|---|---|
| | **All** | Card | Cancer | Mus | Ment | **All** | Card | Cancer | Mus | Ment | **All** | Card | Cancer | Mus | Ment | **All** | Card | Cancer | Mus | Ment |
| Item 1 | **71.1** | 71.1 | 75.6 | 66.3 | 75.6 | **10.9%** | 15.5% | 10.3% | 6.5% | 10.3% | **0.7%** | 1.4% | 0.4% | 0.9% | 0.4% | **0.71** | 0.70 | 0.60 | 0.71 | 0.60 |
| Item 2 | **57.1** | 57.7 | 56.7 | 49.8 | 56.7 | **16.1%** | 24.5% | 17.9% | 9.7% | 17.9% | **1.3%** | 1.8% | 1.1% | 1.4% | 1.1% | **0.66** | 0.67 | 0.65 | 0.72 | 0.65 |
| Item 3 | **75.3** | 75.8 | 82.1 | 70.1 | 82.1 | **19.2%** | 9.7% | 7.0% | 21.2% | 7.0% | **1.1%** | 1.8% | 0.4% | 0.5% | 0.4% | **0.72** | 0.72 | 0.60 | 0.73 | 0.60 |
| Item 4 | **60.9** | 61.9 | 70.0 | 51.5 | 70.0 | **14.5%** | 20.6% | 9.9% | 13.4% | 9.9% | **1.0%** | 1.4% | 0.7% | 0.9% | 0.7% | **0.69** | 0.72 | 0.59 | 0.68 | 0.59 |

**Dimension "Patient involvement in care"**

| | | mean | | | | | standard deviation | | | | |
|---|---|---|---|---|---|---|---|---|---|---|---|
| | Item | **All** | Card | Cancer | Mus | Ment | **All** | Card | Cancer | Mus | Ment |
| Item 1 | I was an equal partner with my healthcare professionals (for example, in making decisions or sharing information). | **5.0** | 5.0 | 5.0 | 4.8 | 5.0 | **1.2** | 1.1 | 1.1 | 1.5 | 1.1 |
| Item 2 | I was informed about various treatment options and their advantages and disadvantages. | **4.3** | 4.4 | 4.5 | 4.0 | 4.5 | **1.7** | 1.7 | 1.6 | 1.9 | 1.6 |
| Item 3 | I was able to participate in the decision-making process as much as I wanted to. | **4.6** | 4.5 | 4.6 | 4.4 | 4.6 | **1.6** | 1.6 | 1.5 | 1.8 | 1.5 |
| Item 4 | When deciding about treatment, it was taken into account what is particularly important to me. | **4.5** | 4.4 | 4.4 | 4.4 | 4.4 | **1.6** | 1.6 | 1.6 | 1.8 | 1.6 |

| | Item difficulty | | | | | 'does not concern me' | | | | | no reply | | | | | item total correlation | | | | |
|---|---|---|---|---|---|---|---|---|---|---|---|---|---|---|---|---|---|---|---|---|
| | **All** | Card | Cancer | Mus | Ment | **All** | Card | Cancer | Mus | Ment | **All** | Card | Cancer | Mus | Ment | **All** | Card | Cancer | Mus | Ment |
| Item 1 | **79.4** | 80.2 | 79.5 | 76.1 | 79.5 | **7.6%** | 13.7% | 5.1% | 6.0% | 5.1% | **1.3%** | 1.4% | 1.1% | 1.8% | 1.1% | **0.66** | 0.71 | 0.62 | 0.77 | 0.62 |
| Item 2 | **65.1** | 68.1 | 69.6 | 59.4 | 69.6 | **27.7%** | 40.8% | 21.2% | 25.3% | 21.2% | **0.9%** | 1.1% | 0.7% | 1.4% | 0.7% | **0.66** | 0.65 | 0.69 | 0.77 | 0.69 |
| Item 3 | **72.0** | 70.7 | 72.5 | 67.1 | 72.5 | **25.2%** | 37.2% | 23.1% | 20.3% | 23.1% | **1.3%** | 1.8% | 1.1% | 0.9% | 1.1% | **0.76** | 0.78 | 0.77 | 0.84 | 0.77 |
| Item 4 | **70.8** | 67.4 | 67.5 | 68.9 | 67.5 | **24.8%** | 40.8% | 24.5% | 17.1% | 24.5% | **1.1%** | 1.8% | 0.7% | 0.9% | 0.7% | **0.79** | 0.82 | 0.79 | 0.88 | 0.79 |

**Dimension "Involvement of family and friends"**

| | Item | mean | | | | | standard deviation | | | | |
|---|---|---|---|---|---|---|---|---|---|---|---|
| | | **All** | Card | Cancer | Mus | Ment | **All** | Card | Cancer | Mus | Ment |
| Item 1 | I was informed about the options for involving my family members in the treatment (for example, accompanying to appointments, participating in conversations, or assisting with medication intake). | **3.1** | 3.0 | 3.4 | 2.2 | 3.4 | **2.0** | 2.0 | 2.0 | 1.7 | 2.0 |
| Item 2 | If I wanted to, my relatives were asked how much they wanted to be involved in my treatment. | **3.0** | 3.0 | 3.5 | 2.3 | 3.5 | **2.0** | 2.1 | 2.0 | 1.9 | 2.0 |
| Item 3 | My relatives were given as much information about my condition and my treatment as I wanted to. | **3.8** | 3.9 | 4.4 | 2.9 | 4.4 | **2.1** | 2.1 | 1.9 | 2.1 | 1.9 |
| Item 4 | My relatives were involved in my treatment as much as I wanted them to be. | **4.0** | 4.1 | 4.4 | 2.9 | 4.4 | **2.1** | 2.0 | 1.9 | 2.1 | 1.9 |

| | Item difficulty | | | | | 'does not concern me' | | | | | no reply | | | | | item total correlation | | | | |
|---|---|---|---|---|---|---|---|---|---|---|---|---|---|---|---|---|---|---|---|---|
| | **All** | Card | Cancer | Mus | Ment | **All** | Card | Cancer | Mus | Ment | **All** | Card | Cancer | Mus | Ment | **All** | Card | Cancer | Mus | Ment |
| Item 1 | **41.5** | 40.4 | 49.0 | 24.4 | 49.0 | **42.6%** | 55.2% | 30.8% | 49.8% | 30.8% | **1.3%** | 1.8% | 2.2% | 0.5% | 2.2% | **0.80** | 0.83 | 0.76 | 0.84 | 0.76 |
| Item 2 | **40.9** | 40.1 | 50.1 | 26.0 | 50.1 | **60.6%** | 67.5% | 49.1% | 67.3% | 49.1% | **1.5%** | 1.8% | 2.9% | 0.5% | 2.9% | **0.85** | 0.84 | 0.82 | 0.90 | 0.82 |
| Item 3 | **56.7** | 58.0 | 68.0 | 38.5 | 68.0 | **57.6%** | 61.7% | 41.8% | 69.6% | 41.8% | **1.5%** | 1.8% | 2.6% | 0.5% | 2.6% | **0.87** | 0.88 | 0.86 | 0.95 | 0.86 |
| Item 4 | **60.0** | 61.5 | 68.8 | 37.7 | 68.8 | **56.4%** | 63.5% | 41.8% | 67.3% | 41.8% | **1.6%** | 1.8% | 3.3% | 0.9% | 3.3% | **0.83** | 0.83 | 0.83 | 0.94 | 0.83 |

**Dimension “Patient empowerment”**

| | Item | mean All | mean Card | mean Cancer | mean Mus | mean Ment | standard deviation All | standard deviation Card | standard deviation Cancer | standard deviation Mus | standard deviation Ment |
|---|---|---|---|---|---|---|---|---|---|---|---|
| Item 1 | I was encouraged to improve my health by changing my behavior (for example, through diet, exercise, reducing tobacco or alcohol). | **4.0** | 3.5 | 3.9 | 3.9 | 3.9 | **1.8** | 1.9 | 1.8 | 1.7 | 1.8 |
| Item 2 | I was encouraged to ask questions. | **4.6** | 4.5 | 4.8 | 4.2 | 4.8 | **1.5** | 1.5 | 1.3 | 1.6 | 1.3 |
| Item 3 | I was explained where to find understandable and scientifically based information about my health. | **3.1** | 2.7 | 3.4 | 2.8 | 3.4 | **1.8** | 1.8 | 1.7 | 1.8 | 1.7 |
| Item 4 | If needed, realistic goals for my health were agreed upon (for example, going for a walk every day, eating fruits every day). | **3.6** | 3.0 | 3.3 | 3.4 | 3.3 | **1.8** | 1.9 | 1.8 | 1.9 | 1.8 |

| | Item difficulty All | Card | Cancer | Mus | Ment | 'does not concern me' All | Card | Cancer | Mus | Ment | no reply All | Card | Cancer | Mus | Ment | item total correlation All | Card | Cancer | Mus | Ment |
|---|---|---|---|---|---|---|---|---|---|---|---|---|---|---|---|---|---|---|---|---|
| Item 1 | **60.9** | 50.7 | 58.6 | 58.9 | 58.6 | **26.0%** | 40.1% | 27.1% | 18.9% | 27.1% | **0.8%** | 1.1% | 1.1% | 0.9% | 1.1% | **0.72** | 0.68 | 0.78 | 0.68 | 0.78 |
| Item 2 | **72.0** | 70.7 | 75.3 | 64.8 | 75.3 | **9.7%** | 16.6% | 8.8% | 6.9% | 8.8% | **1.5%** | 1.4% | 2.9% | 1.4% | 2.9% | **0.57** | 0.55 | 0.53 | 0.66 | 0.53 |
| Item 3 | **42.5** | 34.0 | 47.1 | 36.7 | 47.1 | **21.9%** | 36.8% | 17.2% | 19.4% | 17.2% | **2.0%** | 2.2% | 2.6% | 1.4% | 2.6% | **0.65** | 0.66 | 0.72 | 0.57 | 0.72 |
| Item 4 | **51.1** | 40.6 | 46.9 | 48.9 | 46.9 | **35.7%** | 53.8% | 34.1% | 30.9% | 34.1% | **0.8%** | 1.1% | 1.1% | 0.9% | 1.1% | **0.74** | 0.79 | 0.76 | 0.70 | 0.76 |

**Dimension "Physical support"**

| | Item | mean | | | | | standard deviation | | | | |
|---|---|---|---|---|---|---|---|---|---|---|---|
| | | **All** | Card | Cancer | Mus | Ment | **All** | Card | Cancer | Mus | Ment |
| Item 1 | When I had pain, I was helped quickly. | **4.8** | 5.0 | 5.3 | 4.5 | 5.3 | **1.4** | 1.4 | 0.9 | 1.4 | 0.9 |
| Item 2 | If I had physical complaints, I was helped quickly (for example with nausea or restlessness). | **4.7** | 4.9 | 5.2 | 4.4 | 5.2 | **1.4** | 1.4 | 1.1 | 1.5 | 1.1 |
| Item 3 | I was examined and treated cautiously (for example when giving injections, changing dressings, or washing). | **5.3** | 5.3 | 5.4 | 5.2 | 5.4 | **1.0** | 1.1 | 0.9 | 1.0 | 0.9 |
| Item 4 | If needed, I was asked whether I needed help with everyday tasks (for example, from a care service, home help, or walking frames). | **3.2** | 3.2 | 3.5 | 2.6 | 3.5 | **1.9** | 2.0 | 1.9 | 1.8 | 1.9 |

| | Item difficulty | | | | | 'does not concern me' | | | | | no reply | | | | | item total correlation | | | | |
|---|---|---|---|---|---|---|---|---|---|---|---|---|---|---|---|---|---|---|---|---|
| | **All** | Card | Cancer | Mus | Ment | **All** | Card | Cancer | Mus | Ment | **All** | Card | Cancer | Mus | Ment | **All** | Card | Cancer | Mus | Ment |
| Item 1 | **75.8** | 79.5 | 86.0 | 69.1 | 86.0 | **54.8%** | 69.7% | 52.7% | 19.8% | 52.7% | **1.0%** | 1.1% | 1.8% | 0.9% | 1.8% | **0.66** | 0.61 | 0.61 | 0.64 | 0.61 |
| Item 2 | **75.0** | 77.9 | 83.2 | 68.3 | 83.2 | **56.7%** | 71.1% | 48.0% | 45.6% | 48.0% | **1.2%** | 1.1% | 1.5% | 0.9% | 1.5% | **0.71** | 0.76 | 0.49 | 0.74 | 0.49 |
| Item 3 | **86.6** | 86.0 | 88.8 | 84.6 | 88.8 | **40.4%** | 41.5% | 16.8% | 26.7% | 16.8% | **0.8%** | 0.7% | 1.1% | 0.9% | 1.1% | **0.44** | 0.42 | 0.40 | 0.42 | 0.40 |
| Item 4 | **44.2** | 43.9 | 50.1 | 32.7 | 50.1 | **67.6%** | 78.7% | 59.0% | 59.9% | 59.0% | **0.8%** | 1.1% | 1.1% | 0.9% | 1.1% | **0.41** | 0.47 | 0.41 | 0.41 | 0.41 |

**Dimension "Emotional support"**

| | Item | mean | | | | | standard deviation | | | | |
|---|---|---|---|---|---|---|---|---|---|---|---|
| | | **All** | Card | Cancer | Mus | Ment | **All** | Card | Cancer | Mus | Ment |
| Item 1 | The healthcare professionals addressed my fears and concerns (for example, by showing understanding and providing encouragement). | **4.4** | 3.8 | 4.2 | 3.9 | 4.2 | **1.7** | 1.9 | 1.6 | 1.7 | 1.6 |
| Item 2 | I had the opportunity to talk to my healthcare professionals about my feelings. | **4.4** | 3.9 | 4.2 | 3.9 | 4.2 | **1.7** | 1.8 | 1.6 | 1.7 | 1.6 |
| Item 3 | I was encouraged to talk about my feelings. | **3.8** | 3.2 | 3.4 | 3.1 | 3.4 | **1.9** | 1.8 | 1.8 | 1.8 | 1.8 |
| Item 4 | I was asked whether I would like psychological support (for example, psychological counselling, psychotherapy, or pastoral care). | **3.5** | 2.2 | 3.6 | 2.3 | 3.6 | **2.1** | 1.7 | 2.0 | 1.8 | 2.0 |

| | Item difficulty | | | | | 'does not concern me' | | | | | no reply | | | | | item total correlation | | | | |
|---|---|---|---|---|---|---|---|---|---|---|---|---|---|---|---|---|---|---|---|---|
| | **All** | Card | Cancer | Mus | Ment | **All** | Card | Cancer | Mus | Ment | **All** | Card | Cancer | Mus | Ment | **All** | Card | Cancer | Mus | Ment |
| Item 1 | **67.8** | 56.9 | 63.2 | 58.6 | 63.2 | **19.2%** | 36.1% | 18.7% | 19.4% | 18.7% | **0.8%** | 0.7% | 1.5% | 0.5% | 1.5% | **0.85** | 0.85 | 0.85 | 0.81 | 0.85 |
| Item 2 | **69.0** | 57.0 | 64.6 | 57.5 | 64.6 | **16.8%** | 33.6% | 14.3% | 17.1% | 14.3% | **0.7%** | 1.1% | 1.1% | 0.5% | 1.1% | **0.84** | 0.83 | 0.79 | 0.83 | 0.79 |
| Item 3 | **56.8** | 43.0 | 47.8 | 42.1 | 47.8 | **17.9%** | 33.2% | 16.1% | 19.8% | 16.1% | **1.3%** | 2.2% | 1.5% | 0.9% | 1.5% | **0.87** | 0.85 | 0.83 | 0.86 | 0.83 |
| Item 4 | **49.6** | 23.1 | 52.4 | 26.5 | 52.4 | **31.5%** | 52.3% | 21.6% | 33.2% | 21.6% | **0.9%** | 1.1% | 1.5% | 0.5% | 1.5% | **0.66** | 0.62 | 0.61 | 0.49 | 0.61 |
