## Appendix 5 Item characteristics per medical condition for "Through the patients’ eyes - Psychometric evaluation of the 64-item version of the Experienced Patient-Centeredness Questionnaire (EPAT-64)"

**Appendix 5: Item characteristics per disease group - inpatient sample**

**Article:** Through the patients' eyes - Psychometric evaluation of the Experienced Patient-Centeredness Questionnaire (EPAT-64)

**Authors:** Eva Christalle, Stefan Zeh, Hannah Führes, Alica Schellhorn, Pola Hahlweg, Jördis Zill, Martin Härter, Carsten Bokemeyer, Jürgen Gallinat, Christoffer Gebhardt, Christina Magnussen, Volkmar Müller, Katharina Schmalstieg-Bahr, André Strahl, Levente Kriston, Isabelle Scholl

**Abbreviations:**

- Card = Cardiovascular diseases
- Mus = Musloskeletal diseases
- Ment = Mental disorders

**Dimension „Essential characteristics of the clinicians"**

| | Item | mean | | | | | standard deviation | | | | |
|---|---|---|---|---|---|---|---|---|---|---|---|
| | | **All** | Card | Cancer | Mus | Ment | **All** | Card | Cancer | Mus | Ment |
| Item 1 | The healthcare professionals were sensitive (for example they addressed my feelings, showed understanding, or empathized with my situation). | **5.1** | 5.1 | 5.5 | 4.6 | 5.5 | **1.2** | 1.2 | 0.8 | 1.3 | 0.8 |
| Item 2 | The healthcare professionals behaved respectfully and appreciatively. | **5.5** | 5.6 | 5.8 | 5.2 | 5.8 | **0.9** | 0.9 | 0.6 | 1.1 | 0.6 |
| Item 3 | The healthcare professionals were committed to finding a solution for my health concerns. | **5.3** | 5.4 | 5.6 | 4.9 | 5.6 | **1.0** | 1.0 | 0.7 | 1.1 | 0.7 |
| Item 4 | If I wanted to, difficult topics were discussed directly and openly by the healthcare professionals (for example, long-term effects of the illness, life expectancy, or sexuality). | **4.7** | 4.7 | 5.1 | 4.0 | 5.1 | **1.5** | 1.5 | 1.3 | 1.7 | 1.3 |

| | item difficulty | | | | | 'does not concern me' | | | | | no reply | | | | | item total correlation | | | | |
|---|---|---|---|---|---|---|---|---|---|---|---|---|---|---|---|---|---|---|---|---|
| | **All** | Card | Cancer | Mus | Ment | **All** | Card | Cancer | Mus | Ment | **All** | Card | Cancer | Mus | Ment | **All** | Card | Cancer | Mus | Ment |
| Item 1 | **81.9** | 82.5 | 89.0 | 71.6 | 89.0 | **3.9%** | 7.7% | 2.3% | 5.4% | 2.3% | **0.8%** | 0.7% | 0.9% | 1.1% | 0.9% | **0.73** | 0.61 | 0.67 | 0.76 | 0.67 |
| Item 2 | **89.5** | 91.3 | 95.1 | 84.9 | 95.1 | **0.2%** | 0.0% | 0.0% | 1.1% | 0.0% | **0.9%** | 0.7% | 1.2% | 1.1% | 1.2% | **0.75** | 0.74 | 0.67 | 0.74 | 0.67 |
| Item 3 | **85.0** | 88.1 | 91.3 | 78.7 | 91.3 | **2.9%** | 4.9% | 2.1% | 1.1% | 2.1% | **1.1%** | 1.0% | 1.5% | 0.0% | 1.5% | **0.74** | 0.67 | 0.70 | 0.64 | 0.70 |
| Item 4 | **74.7** | 73.4 | 82.4 | 60.3 | 82.4 | **30.3%** | 39.5% | 27.3% | 33.7% | 27.3% | **1.0%** | 1.4% | 1.2% | 0.0% | 1.2% | **0.65** | 0.51 | 0.64 | 0.70 | 0.64 |

**Dimension „Clinician-patient relationship"**

| | | mean | | | | | standard deviation | | | | |
|---|---|---|---|---|---|---|---|---|---|---|---|
| | Item | **All** | Card | Cancer | Mus | Ment | **All** | Card | Cancer | Mus | Ment |
| Item 1 | I trusted my healthcare professionals. | **5.3** | 5.4 | 5.6 | 4.9 | 5.6 | **1.1** | 1.0 | 0.8 | 1.3 | 0.8 |
| Item 2 | I felt I could confide in my healthcare professionals (for example, on intimate or difficult topics). | **4.9** | 4.8 | 5.2 | 4.4 | 5.2 | **1.3** | 1.4 | 1.1 | 1.7 | 1.1 |
| Item 3 | I was able to talk to the healthcare professionals in a confidential setting (for example, in private, without anyone listening). | **4.8** | 4.4 | 4.8 | 4.5 | 4.8 | **1.6** | 1.7 | 1.6 | 1.6 | 1.6 |
| Item 4 | The healthcare professionals knew about my medical history and my current health status. | **5.2** | 5.3 | 5.4 | 5.0 | 5.4 | **1.0** | 1.0 | 0.8 | 1.1 | 0.8 |

| | item difficulty | | | | | 'does not concern me' | | | | | no reply | | | | | item total correlation | | | | |
|---|---|---|---|---|---|---|---|---|---|---|---|---|---|---|---|---|---|---|---|---|
| | **All** | Card | Cancer | Mus | Ment | **All** | Card | Cancer | Mus | Ment | **All** | Card | Cancer | Mus | Ment | **All** | Card | Cancer | Mus | Ment |
| Item 1 | **86.3** | 88.6 | 91.2 | 78.7 | 91.2 | **1.2%** | 2.4% | 0.3% | 2.2% | 0.3% | **0.4%** | 1.0% | 0.3% | 0.0% | 0.3% | **0.71** | 0.66 | 0.68 | 0.81 | 0.68 |
| Item 2 | **78.6** | 76.8 | 84.1 | 68.3 | 84.1 | **12.9%** | 24.8% | 8.8% | 17.4% | 8.8% | **0.8%** | 1.7% | 0.3% | 1.1% | 0.3% | **0.75** | 0.72 | 0.70 | 0.78 | 0.70 |
| Item 3 | **75.4** | 67.2 | 75.6 | 69.9 | 75.6 | **12.6%** | 23.4% | 12.3% | 3.3% | 12.3% | **0.6%** | 1.0% | 0.3% | 2.2% | 0.3% | **0.52** | 0.58 | 0.59 | 0.55 | 0.59 |
| Item 4 | **84.4** | 85.6 | 88.5 | 79.2 | 88.5 | **1.3%** | 1.7% | 1.2% | 1.1% | 1.2% | **0.5%** | 1.4% | 0.3% | 0.0% | 0.3% | **0.61** | 0.66 | 0.55 | 0.58 | 0.55 |

**Dimension „Patient as a unique person"**

| | | mean | | | | | standard deviation | | | | |
|---|---|---|---|---|---|---|---|---|---|---|---|
| | Item | **All** | Card | Cancer | Mus | Ment | **All** | Card | Cancer | Mus | Ment |
| Item 1 | My wishes, needs and expectations were asked and taken into account in the treatment. | **4.8** | 4.8 | 5.1 | 4.3 | 5.1 | **1.3** | 1.4 | 1.2 | 1.6 | 1.2 |
| Item 2 | My healthcare professionals addressed me personally and did not treat me as just one of many patients. | **5.1** | 5.2 | 5.4 | 4.5 | 5.4 | **1.2** | 1.1 | 0.9 | 1.5 | 0.9 |
| Item 3 | My personal health goals were asked and taken into account. | **4.1** | 3.7 | 4.2 | 3.8 | 4.2 | **1.7** | 1.8 | 1.7 | 1.7 | 1.7 |
| Item 4 | It was asked and taken into account what opportunities and skills I can provide to support my health. | **3.7** | 3.3 | 3.6 | 3.4 | 3.6 | **1.7** | 1.8 | 1.7 | 1.9 | 1.7 |

| | item difficulty | | | | | 'does not concern me' | | | | | no reply | | | | | item total correlation | | | | |
|---|---|---|---|---|---|---|---|---|---|---|---|---|---|---|---|---|---|---|---|---|
| | **All** | Card | Cancer | Mus | Ment | **All** | Card | Cancer | Mus | Ment | **All** | Card | Cancer | Mus | Ment | **All** | Card | Cancer | Mus | Ment |
| Item 1 | **75.8** | 76.9 | 81.5 | 65.2 | 81.5 | **5.5%** | 9.4% | 5.9% | 3.3% | 5.9% | **0.9%** | 1.0% | 0.6% | 0.0% | 0.6% | **0.73** | 0.60 | 0.74 | 0.86 | 0.74 |
| Item 2 | **82.0** | 83.9 | 88.4 | 70.2 | 88.4 | **1.6%** | 2.8% | 1.8% | 0.0% | 1.8% | **0.6%** | 0.7% | 0.3% | 0.0% | 0.3% | **0.58** | 0.49 | 0.55 | 0.75 | 0.55 |
| Item 3 | **62.2** | 54.1 | 64.1 | 56.8 | 64.1 | **12.8%** | 17.8% | 16.4% | 4.3% | 16.4% | **0.9%** | 1.0% | 0.6% | 0.0% | 0.6% | **0.72** | 0.68 | 0.76 | 0.77 | 0.76 |
| Item 4 | **53.9** | 47.0 | 52.9 | 47.8 | 52.9 | **14.5%** | 22.0% | 16.7% | 6.5% | 16.7% | **0.9%** | 0.7% | 0.3% | 1.1% | 0.3% | **0.63** | 0.62 | 0.65 | 0.69 | 0.65 |

**Dimension „Biopsychosocial perspective"**

| | | mean | | | | | standard deviation | | | | |
|---|---|---|---|---|---|---|---|---|---|---|---|
| | Item | **All** | Card | Cancer | Mus | Ment | **All** | Card | Cancer | Mus | Ment |
| Item 1 | My entire personal life was taken into account during the treatment (for example, job, family and friends, partnership and sexuality, culture and religion, age, or financial circumstances). | **3.8** | 3.3 | 3.8 | 3.5 | 3.8 | **1.8** | 1.7 | 1.8 | 1.9 | 1.8 |
| Item 2 | I was asked how my condition affects my life. | **3.8** | 3.5 | 3.6 | 3.9 | 3.6 | **1.9** | 1.9 | 1.9 | 1.9 | 1.9 |
| Item 3 | My entire medical history was asked and taken into account. | **4.9** | 4.9 | 5.0 | 4.8 | 5.0 | **1.4** | 1.4 | 1.3 | 1.4 | 1.3 |
| Item 4 | I was informed about the interaction of physical, psychological, and social factors. | **3.4** | 2.9 | 3.5 | 3.0 | 3.5 | **1.8** | 1.8 | 1.8 | 1.9 | 1.8 |

| | item difficulty | | | | | 'does not concern me' | | | | | no reply | | | | | item total correlation | | | | |
|---|---|---|---|---|---|---|---|---|---|---|---|---|---|---|---|---|---|---|---|---|
| | **All** | Card | Cancer | Mus | Ment | **All** | Card | Cancer | Mus | Ment | **All** | Card | Cancer | Mus | Ment | **All** | Card | Cancer | Mus | Ment |
| Item 1 | **55.8** | 45.8 | 55.3 | 49.1 | 55.3 | **14.2%** | 24.1% | 15.8% | 4.3% | 15.8% | **0.8%** | 0.3% | 0.9% | 2.2% | 0.9% | **0.78** | 0.77 | 0.80 | 0.77 | 0.80 |
| Item 2 | **57.0** | 49.5 | 52.3 | 58.2 | 52.3 | **10.2%** | 14.7% | 12.6% | 4.3% | 12.6% | **1.0%** | 1.0% | 1.5% | 0.0% | 1.5% | **0.71** | 0.70 | 0.73 | 0.74 | 0.73 |
| Item 3 | **78.2** | 78.2 | 80.5 | 76.2 | 80.5 | **4.9%** | 6.3% | 6.2% | 2.2% | 6.2% | **0.6%** | 0.3% | 0.9% | 1.1% | 0.9% | **0.53** | 0.52 | 0.52 | 0.63 | 0.52 |
| Item 4 | **48.5** | 38.0 | 49.1 | 40.5 | 49.1 | **13.0%** | 19.6% | 14.4% | 8.7% | 14.4% | **0.9%** | 0.7% | 0.9% | 1.1% | 0.9% | **0.72** | 0.69 | 0.74 | 0.79 | 0.74 |

**Dimension „Clinician-patient communication"**

| | | mean | | | | | standard deviation | | | | |
|---|---|---|---|---|---|---|---|---|---|---|---|
| | Item | **All** | Card | Cancer | Mus | Ment | **All** | Card | Cancer | Mus | Ment |
| Item 1 | I was given enough time to describe my concerns and my situation (for example, medical history or current symptoms). | **5.2** | 5.3 | 5.5 | 4.8 | 5.5 | **1.1** | 1.0 | 0.9 | 1.3 | 0.9 |
| Item 2 | The healthcare professionals used terms that were easy to understand. | **5.2** | 5.1 | 5.3 | 4.9 | 5.3 | **1.1** | 1.1 | 0.9 | 1.4 | 0.9 |
| Item 3 | The healthcare professionals looked at me and listened carefully during our conversation. | **5.5** | 5.5 | 5.7 | 5.1 | 5.7 | **0.9** | 0.8 | 0.7 | 1.2 | 0.7 |
| Item 4 | The healthcare professionals ensured that I understood correctly what was explained to me. | **5.0** | 5.2 | 5.2 | 4.5 | 5.2 | **1.2** | 1.2 | 1.0 | 1.5 | 1.0 |

| | item difficulty | | | | | 'does not concern me' | | | | | no reply | | | | | item total correlation | | | | |
|---|---|---|---|---|---|---|---|---|---|---|---|---|---|---|---|---|---|---|---|---|
| | **All** | Card | Cancer | Mus | Ment | **All** | Card | Cancer | Mus | Ment | **All** | Card | Cancer | Mus | Ment | **All** | Card | Cancer | Mus | Ment |
| Item 1 | **85.0** | 87.0 | 89.9 | 76.5 | 89.9 | **2.8%** | 2.1% | 5.3% | 0.0% | 5.3% | **0.5%** | 0.3% | 0.9% | 0.0% | 0.9% | **0.72** | 0.71 | 0.69 | 0.81 | 0.69 |
| Item 2 | **83.7** | 81.9 | 86.8 | 78.0 | 86.8 | **0.5%** | 0.7% | 0.6% | 0.0% | 0.6% | **0.5%** | 0.3% | 0.9% | 0.0% | 0.9% | **0.67** | 0.73 | 0.63 | 0.70 | 0.63 |
| Item 3 | **89.6** | 90.7 | 93.2 | 81.5 | 93.2 | **0.2%** | 0.3% | 0.3% | 0.0% | 0.3% | **0.4%** | 0.3% | 0.9% | 0.0% | 0.9% | **0.74** | 0.80 | 0.68 | 0.68 | 0.68 |
| Item 4 | **80.2** | 83.3 | 84.7 | 70.4 | 84.7 | **2.3%** | 3.1% | 2.1% | 0.0% | 2.1% | **0.9%** | 0.7% | 1.8% | 0.0% | 1.8% | **0.71** | 0.72 | 0.72 | 0.67 | 0.72 |

**Dimension „Integration of medical and non-medical care"**

| | Item | mean | | | | | standard deviation | | | | |
|---|---|---|---|---|---|---|---|---|---|---|---|
| | | **All** | Card | Cancer | Mus | Ment | **All** | Card | Cancer | Mus | Ment |
| Item 1 | I was asked if I use or would like to use additional services (for example, support groups, counseling, health courses, complementary and alternative medicine, or spiritual support/pastoral care). | **3.4** | 2.5 | 3.8 | 2.8 | 3.8 | **2.0** | 1.8 | 2.0 | 1.8 | 2.0 |
| Item 2 | If I used or wanted to use additional services, it was accepted. | **4.3** | 3.2 | 4.7 | 3.8 | 4.7 | **1.8** | 1.9 | 1.7 | 2.0 | 1.7 |
| Item 3 | The healthcare professionals informed me about the advantages and disadvantages of additional services. | **3.3** | 2.9 | 3.6 | 2.7 | 3.6 | **1.9** | 1.9 | 2.0 | 2.0 | 2.0 |
| Item 4 | If necessary, I was given specific contacts where I could get information about additional offers. | **3.6** | 2.8 | 4.0 | 3.0 | 4.0 | **2.0** | 1.9 | 2.0 | 2.1 | 2.0 |

| | item difficulty | | | | | 'does not concern me' | | | | | no reply | | | | | item total correlation | | | | |
|---|---|---|---|---|---|---|---|---|---|---|---|---|---|---|---|---|---|---|---|---|
| | **All** | Card | Cancer | Mus | Ment | **All** | Card | Cancer | Mus | Ment | **All** | Card | Cancer | Mus | Ment | **All** | Card | Cancer | Mus | Ment |
| Item 1 | **48.7** | 30.0 | 56.8 | 35.5 | 56.8 | **25.8%** | 43.7% | 21.4% | 21.7% | 21.4% | **1.5%** | 1.0% | 2.3% | 1.1% | 2.3% | **0.81** | 0.85 | 0.82 | 0.86 | 0.82 |
| Item 2 | **65.8** | 43.3 | 74.8 | 55.1 | 74.8 | **56.7%** | 68.9% | 63.9% | 51.1% | 63.9% | **2.6%** | 3.8% | 3.2% | 0.0% | 3.2% | **0.85** | 0.95 | 0.87 | 0.80 | 0.87 |
| Item 3 | **46.1** | 38.3 | 51.5 | 34.8 | 51.5 | **40.9%** | 51.0% | 44.6% | 33.7% | 44.6% | **2.4%** | 2.8% | 3.5% | 0.0% | 3.5% | **0.81** | 0.88 | 0.83 | 0.85 | 0.83 |
| Item 4 | **52.4** | 35.8 | 59.4 | 39.6 | 59.4 | **42.3%** | 53.1% | 47.8% | 40.2% | 47.8% | **2.7%** | 3.8% | 3.2% | 0.0% | 3.2% | **0.84** | 0.90 | 0.88 | 0.83 | 0.88 |

**Dimension „Teamwork and teambuilding"**

| | | mean | | | | | standard deviation | | | | |
|---|---|---|---|---|---|---|---|---|---|---|---|
| | Item | **All** | Card | Cancer | Mus | Ment | **All** | Card | Cancer | Mus | Ment |
| Item 1 | The processes within the team were well organized. | **5.1** | 5.2 | 5.4 | 4.9 | 5.4 | **1.1** | 1.1 | 0.9 | 1.2 | 0.9 |
| Item 2 | The entire care team was responsible and approachable for me. | **5.1** | 5.2 | 5.4 | 4.8 | 5.4 | **1.2** | 1.1 | 0.8 | 1.3 | 0.8 |
| Item 3 | The care team exchanged information about my current health status (for example, everyone was informed about test results). | **5.0** | 5.1 | 5.3 | 4.6 | 5.3 | **1.2** | 1.1 | 0.9 | 1.5 | 0.9 |
| Item 4 | Various healthcare professionals within the care team have given me contradictory information. | **4.8** | 5.0 | 5.2 | 4.2 | 5.2 | **1.6** | 1.5 | 1.3 | 1.7 | 1.3 |

| | item difficulty | | | | | 'does not concern me' | | | | | no reply | | | | | item total correlation | | | | |
|---|---|---|---|---|---|---|---|---|---|---|---|---|---|---|---|---|---|---|---|---|
| | **All** | Card | Cancer | Mus | Ment | **All** | Card | Cancer | Mus | Ment | **All** | Card | Cancer | Mus | Ment | **All** | Card | Cancer | Mus | Ment |
| Item 1 | **81.8** | 83.9 | 88.6 | 78.0 | 88.6 | **1.2%** | 1.4% | 0.3% | 3.3% | 0.3% | **1.0%** | 1.4% | 0.6% | 1.1% | 0.6% | **0.72** | 0.72 | 0.66 | 0.74 | 0.66 |
| Item 2 | **82.2** | 84.4 | 89.0 | 75.3 | 89.0 | **1.4%** | 2.1% | 0.6% | 2.2% | 0.6% | **1.2%** | 2.1% | 0.6% | 1.1% | 0.6% | **0.71** | 0.66 | 0.66 | 0.63 | 0.66 |
| Item 3 | **80.5** | 82.3 | 86.9 | 72.0 | 86.9 | **3.0%** | 4.5% | 2.3% | 3.3% | 2.3% | **1.6%** | 2.1% | 1.8% | 2.2% | 1.8% | **0.70** | 0.67 | 0.68 | 0.72 | 0.68 |
| Item 4 | **75.3** | 79.0 | 83.6 | 63.1 | 83.6 | **7.5%** | 10.1% | 7.3% | 7.6% | 7.3% | **1.2%** | 1.4% | 1.5% | 1.1% | 1.5% | **0.41** | 0.35 | 0.39 | 0.28 | 0.39 |

**Dimension „Access to care”**

| | | mean | | | | | standard deviation | | | | |
|---|---|---|---|---|---|---|---|---|---|---|---|
| | Item | **All** | Card | Cancer | Mus | Ment | **All** | Card | Cancer | Mus | Ment |
| Item 1 | If I wanted to speak to a physician, they were easily accessible. | **4.8** | 4.9 | 5.2 | 4.5 | 5.2 | **1.2** | 1.2 | 1.0 | 1.4 | 1.0 |
| Item 2 | If my inpatient stay was scheduled, I received an appointment in time. | **5.4** | 5.6 | 5.7 | 5.3 | 5.7 | **1.1** | 0.9 | 0.8 | 1.2 | 0.8 |
| Item 3 | If my inpatient stay was scheduled, I could easily get an appointment (for example via phone, mail, or website) | **5.2** | 5.3 | 5.5 | 5.1 | 5.5 | **1.3** | 1.2 | 1.0 | 1.3 | 1.0 |
| Item 4 | If I rang the bell for the nurse, I was helped quickly. | **5.5** | 5.5 | 5.7 | 5.3 | 5.7 | **0.9** | 0.9 | 0.6 | 1.0 | 0.6 |

| | item difficulty | | | | | 'does not concern me' | | | | | no reply | | | | | item total correlation | | | | |
|---|---|---|---|---|---|---|---|---|---|---|---|---|---|---|---|---|---|---|---|---|
| | **All** | Card | Cancer | Mus | Ment | **All** | Card | Cancer | Mus | Ment | **All** | Card | Cancer | Mus | Ment | **All** | Card | Cancer | Mus | Ment |
| Item 1 | **76.2** | 78.0 | 83.5 | 69.5 | 83.5 | **6.6%** | 9.8% | 5.6% | 6.5% | 5.6% | **1.0%** | 1.0% | 1.5% | 0.0% | 1.5% | **0.54** | 0.55 | 0.54 | 0.39 | 0.54 |
| Item 2 | **88.4** | 91.9 | 93.2 | 85.9 | 93.2 | **18.2%** | 22.4% | 13.2% | 19.6% | 13.2% | **0.9%** | 1.4% | 1.2% | 0.0% | 1.2% | **0.61** | 0.46 | 0.63 | 0.43 | 0.63 |
| Item 3 | **83.3** | 86.1 | 89.5 | 82.4 | 89.5 | **30.4%** | 43.4% | 22.9% | 26.1% | 22.9% | **1.9%** | 3.5% | 2.1% | 0.0% | 2.1% | **0.64** | 0.44 | 0.69 | 0.50 | 0.69 |
| Item 4 | **89.0** | 89.7 | 93.3 | 86.0 | 93.3 | **20.0%** | 9.8% | 10.9% | 31.5% | 10.9% | **1.0%** | 1.4% | 1.2% | 0.0% | 1.2% | **0.54** | 0.56 | 0.41 | 0.46 | 0.41 |

**Dimension „Coordination and continuity of care"**

| | Item | mean | | | | | standard deviation | | | | |
|---|---|---|---|---|---|---|---|---|---|---|---|
| | | **All** | Card | Cancer | Mus | Ment | **All** | Card | Cancer | Mus | Ment |
| Item 1 | It was discussed with me whether follow-up appointments would be useful (for example, for aftercare or further treatment). | **5.0** | 5.0 | 5.3 | 4.8 | 5.3 | **1.4** | 1.4 | 1.2 | 1.5 | 1.2 |
| Item 2 | I was explained how long I will approximately have to wait and why. | **4.2** | 4.1 | 4.6 | 4.0 | 4.6 | **1.6** | 1.7 | 1.5 | 1.8 | 1.5 |
| Item 3 | The healthcare professionals took enough time for me. | **5.1** | 5.2 | 5.4 | 4.9 | 5.4 | **1.1** | 1.1 | 0.9 | 1.1 | 0.9 |
| Item 4 | If required, my follow-up appointments were arranged or it was explained how I could arrange follow-up appointments myself (for example for aftercare or further treatment). | **4.8** | 4.8 | 5.3 | 4.4 | 5.3 | **1.4** | 1.5 | 1.1 | 1.6 | 1.1 |

| | item difficulty | | | | | 'does not concern me' | | | | | no reply | | | | | item total correlation | | | | |
|---|---|---|---|---|---|---|---|---|---|---|---|---|---|---|---|---|---|---|---|---|
| | **All** | Card | Cancer | Mus | Ment | **All** | Card | Cancer | Mus | Ment | **All** | Card | Cancer | Mus | Ment | **All** | Card | Cancer | Mus | Ment |
| Item 1 | **80.0** | 79.9 | 86.3 | 75.1 | 86.3 | **7.2%** | 8.4% | 7.3% | 2.2% | 7.3% | **1.0%** | 1.4% | 1.2% | 1.1% | 1.2% | **0.50** | 0.43 | 0.42 | 0.46 | 0.42 |
| Item 2 | **64.7** | 61.3 | 71.5 | 59.3 | 71.5 | **9.0%** | 7.7% | 9.1% | 6.5% | 9.1% | **1.4%** | 1.7% | 1.2% | 1.1% | 1.2% | **0.53** | 0.48 | 0.51 | 0.57 | 0.51 |
| Item 3 | **82.5** | 83.7 | 89.0 | 78.0 | 89.0 | **0.5%** | 1.4% | 0.0% | 1.1% | 0.0% | **0.5%** | 0.7% | 0.6% | 1.1% | 0.6% | **0.65** | 0.56 | 0.58 | 0.66 | 0.58 |
| Item 4 | **76.1** | 75.6 | 85.2 | 68.9 | 85.2 | **6.7%** | 5.6% | 5.3% | 12.0% | 5.3% | **1.7%** | 1.4% | 2.1% | 2.2% | 2.1% | **0.63** | 0.53 | 0.52 | 0.74 | 0.52 |

**Dimension „Patient safety"**

| | Item | mean | | | | | standard deviation | | | | |
|---|---|---|---|---|---|---|---|---|---|---|---|
| | | **All** | Card | Cancer | Mus | Ment | **All** | Card | Cancer | Mus | Ment |
| Item 1 | I was encouraged to speak up if I noticed inconsistencies in my treatment. | **4.1** | 4.1 | 4.3 | 3.7 | 4.3 | **1.7** | 1.7 | 1.8 | 1.8 | 1.8 |
| Item 2 | I was examined thoroughly and carefully. | **5.3** | 5.5 | 5.6 | 5.1 | 5.6 | **1.0** | 0.8 | 0.7 | 1.2 | 0.7 |
| Item 3 | When I was prescribed new medication, I was asked what other medication I am taking and whether I have any intolerances. | **5.2** | 5.2 | 5.4 | 5.0 | 5.4 | **1.3** | 1.2 | 1.1 | 1.5 | 1.1 |
| Item 4 | I was informed about whom to contact if there was an inconsistency in my treatment or if I wanted to file a complaint. | **2.9** | 2.7 | 3.0 | 2.6 | 3.0 | **1.8** | 1.8 | 1.9 | 1.8 | 1.9 |

| | item difficulty | | | | | 'does not concern me' | | | | | no reply | | | | | item total correlation | | | | |
|---|---|---|---|---|---|---|---|---|---|---|---|---|---|---|---|---|---|---|---|---|
| | **All** | Card | Cancer | Mus | Ment | **All** | Card | Cancer | Mus | Ment | **All** | Card | Cancer | Mus | Ment | **All** | Card | Cancer | Mus | Ment |
| Item 1 | **61.9** | 62.0 | 65.5 | 53.5 | 65.5 | **14.2%** | 15.0% | 19.1% | 8.7% | 19.1% | **1.2%** | 1.0% | 1.5% | 1.1% | 1.5% | **0.65** | 0.63 | 0.65 | 0.66 | 0.65 |
| Item 2 | **86.9** | 90.8 | 92.8 | 83.0 | 92.8 | **1.0%** | 0.3% | 0.9% | 1.1% | 0.9% | **2.3%** | 1.4% | 3.2% | 4.3% | 3.2% | **0.47** | 0.41 | 0.39 | 0.56 | 0.39 |
| Item 3 | **84.1** | 84.9 | 88.7 | 80.7 | 88.7 | **13.1%** | 9.8% | 15.0% | 8.7% | 15.0% | **1.1%** | 0.3% | 1.2% | 2.2% | 1.2% | **0.50** | 0.49 | 0.40 | 0.59 | 0.40 |
| Item 4 | **37.8** | 33.8 | 40.5 | 31.9 | 40.5 | **17.1%** | 17.8% | 22.0% | 13.0% | 22.0% | **1.3%** | 1.4% | 1.5% | 1.1% | 1.5% | **0.49** | 0.47 | 0.44 | 0.53 | 0.44 |

**Dimension „Patient information”**

| | Item | mean | | | | | standard deviation | | | | |
|---|---|---|---|---|---|---|---|---|---|---|---|
| | | **All** | Card | Cancer | Mus | Ment | **All** | Card | Cancer | Mus | Ment |
| Item 1 | I received information about my condition from my healthcare professionals (for example, causes, symptoms, effects or course). | **4.6** | 4.8 | 4.7 | 4.4 | 4.7 | **1.5** | 1.4 | 1.4 | 1.7 | 1.4 |
| Item 2 | I was asked what I already know about my condition. | **4.1** | 4.3 | 4.2 | 3.9 | 4.2 | **1.8** | 1.7 | 1.7 | 1.9 | 1.7 |
| Item 3 | The significance of my test results was explained to me. | **4.8** | 4.8 | 5.1 | 4.6 | 5.1 | **1.5** | 1.4 | 1.2 | 1.7 | 1.2 |
| Item 4 | I was asked what I would like to know about my condition. | **4.2** | 4.4 | 4.4 | 3.9 | 4.4 | **1.8** | 1.7 | 1.7 | 1.9 | 1.7 |

| | item difficulty | | | | | 'does not concern me' | | | | | no reply | | | | | item total correlation | | | | |
|---|---|---|---|---|---|---|---|---|---|---|---|---|---|---|---|---|---|---|---|---|
| | **All** | Card | Cancer | Mus | Ment | **All** | Card | Cancer | Mus | Ment | **All** | Card | Cancer | Mus | Ment | **All** | Card | Cancer | Mus | Ment |
| Item 1 | **72.1** | 76.6 | 73.7 | 68.1 | 73.7 | **6.4%** | 5.2% | 7.9% | 5.4% | 7.9% | **1.3%** | 1.4% | 1.8% | 1.1% | 1.8% | **0.72** | 0.72 | 0.68 | 0.77 | 0.68 |
| Item 2 | **62.3** | 65.7 | 63.6 | 58.9 | 63.6 | **6.6%** | 8.0% | 7.0% | 5.4% | 7.0% | **0.8%** | 1.7% | 0.6% | 0.0% | 0.6% | **0.65** | 0.66 | 0.63 | 0.67 | 0.63 |
| Item 3 | **75.1** | 76.9 | 82.9 | 71.0 | 82.9 | **4.6%** | 2.1% | 4.7% | 3.3% | 4.7% | **1.2%** | 1.4% | 1.8% | 0.0% | 1.8% | **0.63** | 0.64 | 0.58 | 0.72 | 0.58 |
| Item 4 | **63.0** | 68.2 | 68.9 | 57.2 | 68.9 | **8.9%** | 10.8% | 8.8% | 6.5% | 8.8% | **1.4%** | 1.7% | 1.8% | 0.0% | 1.8% | **0.68** | 0.61 | 0.63 | 0.77 | 0.63 |

**Dimension „Patient involvement in care”**

| | | mean | | | | | standard deviation | | | | |
|---|---|---|---|---|---|---|---|---|---|---|---|
| | Item | **All** | Card | Cancer | Mus | Ment | **All** | Card | Cancer | Mus | Ment |
| Item 1 | I was an equal partner with my healthcare professionals (for example, in making decisions or sharing information). | **4.8** | 4.8 | 5.2 | 4.4 | 5.2 | **1.3** | 1.3 | 1.0 | 1.5 | 1.0 |
| Item 2 | I was informed about various treatment options and their advantages and disadvantages. | **4.4** | 4.3 | 5.0 | 4.1 | 5.0 | **1.7** | 1.7 | 1.4 | 1.8 | 1.4 |
| Item 3 | I was able to participate in the decision-making process as much as I wanted to. | **4.3** | 4.1 | 4.7 | 4.0 | 4.7 | **1.7** | 1.7 | 1.6 | 1.8 | 1.6 |
| Item 4 | When deciding about treatment, it was taken into account what is particularly important to me. | **4.3** | 4.0 | 4.7 | 4.1 | 4.7 | **1.6** | 1.8 | 1.5 | 1.7 | 1.5 |

| | item difficulty | | | | | 'does not concern me' | | | | | no reply | | | | | item total correlation | | | | |
|---|---|---|---|---|---|---|---|---|---|---|---|---|---|---|---|---|---|---|---|---|
| | **All** | Card | Cancer | Mus | Ment | **All** | Card | Cancer | Mus | Ment | **All** | Card | Cancer | Mus | Ment | **All** | Card | Cancer | Mus | Ment |
| Item 1 | **75.8** | 75.1 | 83.6 | 68.5 | 83.6 | **4.4%** | 7.3% | 3.8% | 2.2% | 3.8% | **1.8%** | 2.1% | 2.6% | 1.1% | 2.6% | **0.66** | 0.51 | 0.64 | 0.76 | 0.64 |
| Item 2 | **68.8** | 66.7 | 80.4 | 62.7 | 80.4 | **16.4%** | 21.0% | 15.8% | 10.9% | 15.8% | **0.8%** | 0.0% | 1.5% | 1.1% | 1.5% | **0.70** | 0.67 | 0.69 | 0.68 | 0.69 |
| Item 3 | **65.6** | 62.2 | 74.1 | 60.2 | 74.1 | **18.6%** | 25.9% | 19.1% | 7.6% | 19.1% | **1.5%** | 0.7% | 2.9% | 1.1% | 2.9% | **0.78** | 0.74 | 0.82 | 0.80 | 0.82 |
| Item 4 | **66.9** | 60.0 | 74.3 | 62.6 | 74.3 | **18.5%** | 26.6% | 21.7% | 7.6% | 21.7% | **1.7%** | 2.1% | 2.3% | 1.1% | 2.3% | **0.76** | 0.77 | 0.78 | 0.73 | 0.78 |

**Dimension „Involvement of family and friends"**

| | | mean | | | | | standard deviation | | | | |
|---|---|---|---|---|---|---|---|---|---|---|---|
| | Item | **All** | Card | Cancer | Mus | Ment | **All** | Card | Cancer | Mus | Ment |
| Item 1 | I was informed about the options for involving my family members in the treatment (for example, accompanying to appointments, participating in conversations, or assisting with medication intake). | **3.3** | 3.3 | 3.4 | 2.7 | 3.4 | **1.9** | 2.0 | 1.9 | 1.8 | 1.9 |
| Item 2 | If I wanted to, my relatives were asked how much they wanted to be involved in my treatment. | **3.1** | 3.3 | 3.2 | 2.8 | 3.2 | **1.9** | 2.0 | 1.9 | 1.9 | 1.9 |
| Item 3 | My relatives were given as much information about my condition and my treatment as I wanted to. | **4.1** | 4.3 | 4.5 | 3.4 | 4.5 | **1.9** | 1.9 | 1.8 | 1.9 | 1.8 |
| Item 4 | My relatives were involved in my treatment as much as I wanted them to be. | **4.0** | 4.1 | 4.2 | 3.4 | 4.2 | **1.9** | 2.0 | 1.9 | 2.0 | 1.9 |

| | item difficulty | | | | | 'does not concern me' | | | | | no reply | | | | | item total correlation | | | | |
|---|---|---|---|---|---|---|---|---|---|---|---|---|---|---|---|---|---|---|---|---|
| | **All** | Card | Cancer | Mus | Ment | **All** | Card | Cancer | Mus | Ment | **All** | Card | Cancer | Mus | Ment | **All** | Card | Cancer | Mus | Ment |
| Item 1 | **46.0** | 45.4 | 48.7 | 33.0 | 48.7 | **29.6%** | 39.2% | 30.5% | 28.3% | 30.5% | **1.0%** | 1.0% | 1.2% | 0.0% | 1.2% | **0.75** | 0.74 | 0.76 | 0.77 | 0.76 |
| Item 2 | **42.8** | 45.6 | 43.5 | 35.0 | 43.5 | **50.2%** | 53.8% | 50.4% | 43.5% | 50.4% | **1.4%** | 2.1% | 1.5% | 0.0% | 1.5% | **0.82** | 0.87 | 0.79 | 0.78 | 0.79 |
| Item 3 | **62.4** | 65.2 | 70.2 | 47.3 | 70.2 | **40.3%** | 46.2% | 37.0% | 40.2% | 37.0% | **1.4%** | 1.4% | 1.5% | 0.0% | 1.5% | **0.80** | 0.81 | 0.77 | 0.77 | 0.77 |
| Item 4 | **59.5** | 61.3 | 64.2 | 48.0 | 64.2 | **43.0%** | 50.3% | 41.1% | 40.2% | 41.1% | **1.9%** | 2.1% | 2.3% | 0.0% | 2.3% | **0.85** | 0.86 | 0.88 | 0.82 | 0.88 |

**Dimension „Patient empowerment"**

| | | mean | | | | | standard deviation | | | | |
|---|---|---|---|---|---|---|---|---|---|---|---|
| | Item | **All** | Card | Cancer | Mus | Ment | **All** | Card | Cancer | Mus | Ment |
| Item 1 | I was encouraged to improve my health by changing my behavior (for example, through diet, exercise, reducing tobacco or alcohol). | **4.0** | 3.8 | 3.8 | 3.6 | 3.8 | **1.7** | 1.8 | 1.8 | 1.7 | 1.8 |
| Item 2 | I was encouraged to ask questions. | **4.8** | 4.8 | 5.0 | 4.2 | 5.0 | **1.4** | 1.4 | 1.2 | 1.6 | 1.2 |
| Item 3 | I was explained where to find understandable and scientifically based information about my health. | **3.3** | 3.0 | 3.6 | 3.1 | 3.6 | **1.7** | 1.7 | 1.7 | 1.8 | 1.7 |
| Item 4 | If needed, realistic goals for my health were agreed upon (for example, going for a walk every day, eating fruits every day). | **3.7** | 3.4 | 3.7 | 3.4 | 3.7 | **1.8** | 1.9 | 1.8 | 1.7 | 1.8 |

| | item difficulty | | | | | 'does not concern me' | | | | | no reply | | | | | item total correlation | | | | |
|---|---|---|---|---|---|---|---|---|---|---|---|---|---|---|---|---|---|---|---|---|
| | **All** | Card | Cancer | Mus | Ment | **All** | Card | Cancer | Mus | Ment | **All** | Card | Cancer | Mus | Ment | **All** | Card | Cancer | Mus | Ment |
| Item 1 | **60.8** | 56.0 | 56.5 | 52.6 | 56.5 | **25.8%** | 29.0% | 34.0% | 20.7% | 34.0% | **0.5%** | 0.3% | 1.2% | 0.0% | 1.2% | **0.67** | 0.71 | 0.70 | 0.67 | 0.70 |
| Item 2 | **75.0** | 76.8 | 79.8 | 63.8 | 79.8 | **4.8%** | 8.0% | 4.7% | 2.2% | 4.7% | **1.1%** | 0.7% | 1.8% | 1.1% | 1.8% | **0.47** | 0.43 | 0.50 | 0.47 | 0.50 |
| Item 3 | **46.3** | 40.8 | 51.5 | 42.4 | 51.5 | **16.6%** | 24.5% | 15.5% | 10.9% | 15.5% | **1.5%** | 1.4% | 2.1% | 0.0% | 2.1% | **0.61** | 0.58 | 0.66 | 0.60 | 0.66 |
| Item 4 | **54.7** | 48.6 | 53.8 | 48.8 | 53.8 | **29.1%** | 34.6% | 34.9% | 25.0% | 34.9% | **1.0%** | 0.7% | 1.8% | 1.1% | 1.8% | **0.73** | 0.73 | 0.76 | 0.66 | 0.76 |

**Dimension „Physical support”**

| | | mean | | | | | standard deviation | | | | |
|---|---|---|---|---|---|---|---|---|---|---|---|
| | Item | **All** | Card | Cancer | Mus | Ment | **All** | Card | Cancer | Mus | Ment |
| Item 1 | When I had pain, I was helped quickly. | **5.4** | 5.6 | 5.7 | 4.9 | 5.7 | **1.0** | 0.8 | 0.7 | 1.3 | 0.7 |
| Item 2 | If I had physical complaints, I was helped quickly (for example with nausea or restlessness). | **5.2** | 5.4 | 5.6 | 4.8 | 5.6 | **1.1** | 0.9 | 0.8 | 1.3 | 0.8 |
| Item 3 | I was examined and treated cautiously (for example when giving injections, changing dressings, or washing). | **5.5** | 5.5 | 5.7 | 5.0 | 5.7 | **0.9** | 0.9 | 0.6 | 1.3 | 0.6 |
| Item 4 | If needed, I was asked whether I needed help with everyday tasks (for example, from a care service, home help, or walking frames). | **4.2** | 4.2 | 4.5 | 4.2 | 4.5 | **1.8** | 1.9 | 1.8 | 1.9 | 1.8 |

| | item difficulty | | | | | 'does not concern me' | | | | | no reply | | | | | item total correlation | | | | |
|---|---|---|---|---|---|---|---|---|---|---|---|---|---|---|---|---|---|---|---|---|
| | **All** | Card | Cancer | Mus | Ment | **All** | Card | Cancer | Mus | Ment | **All** | Card | Cancer | Mus | Ment | **All** | Card | Cancer | Mus | Ment |
| Item 1 | **87.1** | 91.3 | 94.0 | 77.0 | 94.0 | **19.4%** | 23.8% | 17.0% | 5.4% | 17.0% | **0.8%** | 1.4% | 0.6% | 0.0% | 0.6% | **0.71** | 0.70 | 0.54 | 0.67 | 0.54 |
| Item 2 | **84.9** | 88.4 | 92.2 | 76.3 | 92.2 | **23.8%** | 28.3% | 23.8% | 22.8% | 23.8% | **0.5%** | 0.7% | 0.6% | 0.0% | 0.6% | **0.71** | 0.60 | 0.56 | 0.74 | 0.56 |
| Item 3 | **89.1** | 89.5 | 94.2 | 80.7 | 94.2 | **5.8%** | 2.1% | 1.8% | 12.0% | 1.8% | **0.3%** | 0.0% | 0.6% | 0.0% | 0.6% | **0.57** | 0.47 | 0.46 | 0.51 | 0.46 |
| Item 4 | **64.4** | 63.2 | 70.5 | 64.3 | 70.5 | **48.2%** | 51.7% | 48.7% | 28.3% | 48.7% | **0.6%** | 0.3% | 0.6% | 1.1% | 0.6% | **0.45** | 0.44 | 0.21 | 0.64 | 0.21 |

**Dimension „Emotional support"**

| | Item | mean | | | | | standard deviation | | | | |
|---|---|---|---|---|---|---|---|---|---|---|---|
| | | **All** | Card | Cancer | Mus | Ment | **All** | Card | Cancer | Mus | Ment |
| Item 1 | The healthcare professionals addressed my fears and concerns (for example, by showing understanding and providing encouragement). | **4.4** | 4.1 | 4.7 | 3.6 | 4.7 | **1.6** | 1.7 | 1.4 | 1.8 | 1.4 |
| Item 2 | I had the opportunity to talk to my healthcare professionals about my feelings. | **4.3** | 3.8 | 4.5 | 3.2 | 4.5 | **1.7** | 1.8 | 1.6 | 1.7 | 1.6 |
| Item 3 | I was encouraged to talk about my feelings. | **3.7** | 3.1 | 3.5 | 2.6 | 3.5 | **1.8** | 1.7 | 1.8 | 1.7 | 1.8 |
| Item 4 | I was asked whether I would like psychological support (for example, psychological counselling, psychotherapy, or pastoral care). | **3.9** | 2.6 | 4.7 | 2.3 | 4.7 | **2.0** | 1.8 | 1.8 | 1.8 | 1.8 |

| | item difficulty | | | | | 'does not concern me' | | | | | no reply | | | | | item total correlation | | | | |
|---|---|---|---|---|---|---|---|---|---|---|---|---|---|---|---|---|---|---|---|---|
| | **All** | Card | Cancer | Mus | Ment | **All** | Card | Cancer | Mus | Ment | **All** | Card | Cancer | Mus | Ment | **All** | Card | Cancer | Mus | Ment |
| Item 1 | **68.8** | 62.3 | 74.5 | 51.3 | 74.5 | **15.9%** | 25.2% | 17.9% | 14.1% | 17.9% | **1.5%** | 0.7% | 2.6% | 1.1% | 2.6% | **0.76** | 0.74 | 0.73 | 0.76 | 0.73 |
| Item 2 | **66.4** | 55.4 | 69.9 | 43.6 | 69.9 | **18.3%** | 28.7% | 20.5% | 18.5% | 20.5% | **1.4%** | 1.7% | 1.2% | 2.2% | 1.2% | **0.84** | 0.87 | 0.76 | 0.82 | 0.76 |
| Item 3 | **53.7** | 41.3 | 50.7 | 31.1 | 50.7 | **19.5%** | 30.4% | 21.1% | 20.7% | 21.1% | **1.7%** | 1.4% | 2.3% | 1.1% | 2.3% | **0.79** | 0.86 | 0.72 | 0.78 | 0.72 |
| Item 4 | **57.9** | 31.2 | 74.6 | 25.9 | 74.6 | **23.4%** | 39.2% | 13.5% | 25.0% | 13.5% | **1.0%** | 0.7% | 1.2% | 1.1% | 1.2% | **0.63** | 0.59 | 0.48 | 0.69 | 0.48 |
