## Appendix 6 Correlations dimensions for "Through the patients’ eyes - Psychometric evaluation of the 64-item version of the Experienced Patient-Centeredness Questionnaire (EPAT-64)"

**Appendix 6: Correlations between specific dimensions**

**Article:** Through the patients' eyes - Psychometric evaluation of the Experienced Patient-Centeredness Questionnaire (EPAT-64)

**Authors:** Eva Christalle, Stefan Zeh, Hannah Führes, Alica Schellhorn, Pola Hahlweg, Jördis Zill, Martin Härter, Carsten Bokemeyer, Jürgen Gallinat, Christoffer Gebhardt, Christina Magnussen, Volkmar Müller, Katharina Schmalstieg-Bahr, André Strahl, Levente Kriston, Isabelle Scholl

**Table 1:** Correlations between specific dimensions estimated based on confirmatory factor analysis - outpatient sample

| | Patient as a unique person | Patient involvement in care | Physical support | Patient empowerment | Clinician-patient communication | Patient safety | Patient information | Involvement of family and friends | Teamwork and teambuilding | Clinician-patient relationship | Biopsychosocial perspective | Emotional support | Coordination and continuity of care | Essential characteristics of the clinicians | Access to care | Integration of medical and non-medical care |
|---|---|---|---|---|---|---|---|---|---|---|---|---|---|---|---|---|
| Patient as a unique person | 1.000 | 0.703 | 0.278 | 0.824 | 0.627 | 0.734 | 0.643 | 0.604 | 0.279 | 0.580 | 0.800 | 0.712 | 0.700 | 0.661 | 0.136 | 0.630 |
| Patient involvement in care | 0.703 | 1.000 | 0.341 | 0.672 | 0.447 | 0.527 | 0.509 | 0.520 | 0.191 | 0.416 | 0.604 | 0.522 | 0.580 | 0.426 | 0.150 | 0.521 |
| Physical support | 0.278 | 0.341 | 1.000 | 0.450 | 0.228 | 0.415 | 0.258 | 0.457 | 0.304 | 0.323 | 0.323 | 0.258 | 0.344 | 0.364 | 0.281 | 0.361 |
| Patient empowerment | 0.824 | 0.672 | 0.450 | 1.000 | 0.658 | 0.765 | 0.725 | 0.691 | 0.370 | 0.580 | 0.736 | 0.645 | 0.780 | 0.647 | 0.226 | 0.724 |
| Clinician-patient communication | 0.627 | 0.447 | 0.228 | 0.658 | 1.000 | 0.605 | 0.443 | 0.478 | 0.235 | 0.453 | 0.566 | 0.559 | 0.602 | 0.471 | 0.269 | 0.528 |
| Patient safety | 0.734 | 0.527 | 0.415 | 0.765 | 0.605 | 1.000 | 0.676 | 0.708 | 0.370 | 0.476 | 0.710 | 0.524 | 0.782 | 0.579 | 0.256 | 0.625 |
| Patient information | 0.643 | 0.509 | 0.258 | 0.725 | 0.443 | 0.676 | 1.000 | 0.614 | 0.353 | 0.494 | 0.619 | 0.436 | 0.750 | 0.515 | 0.214 | 0.522 |
| Involvement of family and friends | 0.604 | 0.520 | 0.457 | 0.691 | 0.478 | 0.708 | 0.614 | 1.000 | 0.416 | 0.528 | 0.599 | 0.489 | 0.710 | 0.525 | 0.302 | 0.596 |
| Teamwork and teambuilding | 0.279 | 0.191 | 0.304 | 0.370 | 0.235 | 0.370 | 0.353 | 0.416 | 1.000 | 0.260 | 0.299 | 0.224 | 0.605 | 0.236 | 0.517 | 0.336 |
| Clinician-patient relationship | 0.580 | 0.416 | 0.323 | 0.580 | 0.453 | 0.476 | 0.494 | 0.528 | 0.260 | 1.000 | 0.564 | 0.586 | 0.445 | 0.660 | 0.040 | 0.487 |
| Biopsychosocial perspective | 0.800 | 0.604 | 0.323 | 0.736 | 0.566 | 0.710 | 0.619 | 0.599 | 0.299 | 0.564 | 1.000 | 0.776 | 0.714 | 0.629 | 0.123 | 0.653 |
| Emotional support | 0.712 | 0.522 | 0.258 | 0.645 | 0.559 | 0.524 | 0.436 | 0.489 | 0.224 | 0.586 | 0.776 | 1.000 | 0.606 | 0.697 | 0.109 | 0.622 |
| Coordination and continuity of care | 0.700 | 0.580 | 0.344 | 0.780 | 0.602 | 0.782 | 0.750 | 0.710 | 0.605 | 0.445 | 0.714 | 0.606 | 1.000 | 0.568 | 0.489 | 0.636 |
| Essential characteristics of the clinicians | 0.661 | 0.426 | 0.364 | 0.647 | 0.471 | 0.579 | 0.515 | 0.525 | 0.236 | 0.660 | 0.629 | 0.697 | 0.568 | 1.000 | 0.146 | 0.551 |
| Access to care | 0.136 | 0.150 | 0.281 | 0.226 | 0.269 | 0.256 | 0.214 | 0.302 | 0.517 | 0.040 | 0.123 | 0.109 | 0.489 | 0.146 | 1.000 | 0.239 |
| Integration of medical and non-medical care | 0.630 | 0.521 | 0.361 | 0.724 | 0.528 | 0.625 | 0.522 | 0.596 | 0.336 | 0.487 | 0.653 | 0.622 | 0.636 | 0.551 | 0.239 | 1.000 |

**Table 2:** Correlations between specific dimensions estimated based on confirmatory factor analysis - inpatient sample

| | Patient as a unique person | Patient involvement in care | Physical support | Patient empowerment | Clinician-patient communication | Patient safety | Patient information | Involvement of family and friends | Teamwork and teambuilding | Clinician-patient relationship | Biopsychosocial perspective | Emotional support | Coordination and continuity of care | Essential characteristics of the clinicians | Access to care | Integration of medical and non-medical care |
|---|---|---|---|---|---|---|---|---|---|---|---|---|---|---|---|---|
| Patient as a unique person | 1.000 | 0.648 | 0.098 | 0.773 | 0.545 | 0.707 | 0.589 | 0.610 | 0.236 | 0.614 | 0.813 | 0.664 | 0.655 | 0.596 | -0.041 | 0.655 |
| Patient involvement in care | 0.648 | 1.000 | 0.019 | 0.505 | 0.364 | 0.488 | 0.470 | 0.478 | 0.107 | 0.420 | 0.525 | 0.435 | 0.523 | 0.383 | -0.021 | 0.508 |
| Physical support | 0.098 | 0.019 | 1.000 | 0.094 | -0.045 | 0.118 | 0.036 | 0.107 | 0.063 | -0.046 | 0.060 | 0.073 | 0.040 | 0.054 | 0.231 | 0.137 |
| Patient empowerment | 0.773 | 0.505 | 0.094 | 1.000 | 0.456 | 0.735 | 0.595 | 0.582 | 0.308 | 0.518 | 0.694 | 0.595 | 0.650 | 0.525 | -0.003 | 0.645 |
| Clinician-patient communication | 0.545 | 0.364 | -0.045 | 0.456 | 1.000 | 0.384 | 0.345 | 0.288 | 0.169 | 0.465 | 0.478 | 0.490 | 0.329 | 0.265 | -0.070 | 0.353 |
| Patient safety | 0.707 | 0.488 | 0.118 | 0.735 | 0.384 | 1.000 | 0.594 | 0.631 | 0.324 | 0.415 | 0.658 | 0.482 | 0.697 | 0.446 | 0.021 | 0.578 |
| Patient information | 0.589 | 0.470 | 0.036 | 0.595 | 0.345 | 0.594 | 1.000 | 0.545 | 0.314 | 0.360 | 0.600 | 0.391 | 0.641 | 0.300 | 0.004 | 0.421 |
| Involvement of family and friends | 0.610 | 0.478 | 0.107 | 0.582 | 0.288 | 0.631 | 0.545 | 1.000 | 0.314 | 0.369 | 0.595 | 0.482 | 0.678 | 0.436 | 0.031 | 0.594 |
| Teamwork and teambuilding | 0.236 | 0.107 | 0.063 | 0.308 | 0.169 | 0.324 | 0.314 | 0.314 | 1.000 | 0.204 | 0.278 | 0.235 | 0.363 | 0.059 | -0.140 | 0.280 |
| Clinician-patient relationship | 0.614 | 0.420 | -0.046 | 0.518 | 0.465 | 0.415 | 0.360 | 0.369 | 0.204 | 1.000 | 0.599 | 0.667 | 0.427 | 0.523 | -0.114 | 0.471 |
| Biopsychosocial perspective | 0.813 | 0.525 | 0.060 | 0.694 | 0.478 | 0.658 | 0.600 | 0.595 | 0.278 | 0.599 | 1.000 | 0.744 | 0.740 | 0.589 | 0.000 | 0.656 |
| Emotional support | 0.664 | 0.435 | 0.073 | 0.595 | 0.490 | 0.482 | 0.391 | 0.482 | 0.235 | 0.667 | 0.744 | 1.000 | 0.621 | 0.719 | 0.020 | 0.626 |
| Coordination and continuity of care | 0.655 | 0.523 | 0.040 | 0.650 | 0.329 | 0.697 | 0.641 | 0.678 | 0.363 | 0.427 | 0.740 | 0.621 | 1.000 | 0.580 | 0.131 | 0.673 |
| Essential characteristics of the clinicians | 0.596 | 0.383 | 0.054 | 0.525 | 0.265 | 0.446 | 0.300 | 0.436 | 0.059 | 0.523 | 0.589 | 0.719 | 0.580 | 1.000 | -0.051 | 0.553 |
| Access to care | -0.041 | -0.021 | 0.231 | -0.003 | -0.070 | 0.021 | 0.004 | 0.031 | -0.140 | -0.114 | 0.000 | 0.020 | 0.131 | -0.051 | 1.000 | 0.085 |
| Integration of medical and non-medical care | 0.655 | 0.508 | 0.137 | 0.645 | 0.353 | 0.578 | 0.421 | 0.594 | 0.280 | 0.471 | 0.656 | 0.626 | 0.673 | 0.553 | 0.085 | 1.000 |
