## Appendix 7 Correlations sum scores for "Through the patients’ eyes - Psychometric evaluation of the 64-item version of the Experienced Patient-Centeredness Questionnaire (EPAT-64)"

**Appendix 7: Correlations between sum scores and model-based factor scores**

**Article:** Through the patients' eyes - Psychometric evaluation of the Experienced Patient-Centeredness Questionnaire (EPAT-64)

**Authors:** Eva Christalle, Stefan Zeh, Hannah Führes, Alica Schellhorn, Pola Hahlweg, Jördis Zill, Martin Härter, Carsten Bokemeyer, Jürgen Gallinat, Christoffer Gebhardt, Christina Magnussen, Volkmar Müller, Katharina Schmalstieg-Bahr, André Strahl, Levente Kriston, Isabelle Scholl

**Table 1:** Correlations between sum scores and model-based factor scores - outpatient sample

| Model-based factor scores / Sum scores for specific dimensions | Essential characteristics of the clinicians | Clinician-patient relationship | Patient as a unique person | Biopsychosocial perspective | Clinician-patient communication | Integration of medical and non-medical | Teamwork and teambuilding | Access to care | Coordination and continuity of care | Patient safety | Patient information | Patient involvement in care | Involvement of family and friends | Patient empowerment | Physical support | Emotional support | General factor |
|---|---|---|---|---|---|---|---|---|---|---|---|---|---|---|---|---|---|
| Essential characteristics of the clinicians | **0.560** | 0.368 | 0.324 | 0.303 | 0.267 | 0.280 | 0.127 | 0.085 | 0.289 | 0.301 | 0.267 | 0.213 | 0.274 | 0.320 | 0.219 | 0.335 | 0.872 |
| Clinician-patient relationship | 0.428 | **0.588** | 0.360 | 0.355 | 0.314 | 0.318 | 0.211 | 0.070 | 0.321 | 0.334 | 0.340 | 0.280 | 0.353 | 0.374 | 0.242 | 0.343 | 0.823 |
| Patient as a unique person | 0.602 | 0.538 | **0.802** | 0.643 | 0.577 | 0.530 | 0.230 | 0.106 | 0.580 | 0.626 | 0.533 | 0.600 | 0.509 | 0.665 | 0.263 | 0.574 | 0.628 |
| Biopsychosocial perspective | 0.636 | 0.558 | 0.725 | **0.895** | 0.582 | 0.606 | 0.277 | 0.097 | 0.665 | 0.682 | 0.594 | 0.563 | 0.562 | 0.671 | 0.338 | 0.684 | 0.423 |
| Clinician-patient communication | 0.296 | 0.289 | 0.354 | 0.315 | **0.632** | 0.317 | 0.154 | 0.167 | 0.348 | 0.356 | 0.254 | 0.271 | 0.283 | 0.368 | 0.161 | 0.311 | 0.798 |
| Integration of medical and non-medical care | 0.608 | 0.538 | 0.622 | 0.645 | 0.609 | **0.970** | 0.305 | 0.207 | 0.629 | 0.641 | 0.506 | 0.530 | 0.597 | 0.713 | 0.407 | 0.619 | 0.286 |
| Teamwork and teambuilding | 0.172 | 0.204 | 0.185 | 0.195 | 0.173 | 0.227 | **0.730** | 0.371 | 0.419 | 0.249 | 0.248 | 0.139 | 0.298 | 0.243 | 0.232 | 0.149 | 0.609 |
| Access to care | 0.175 | 0.049 | 0.146 | 0.135 | 0.275 | 0.227 | 0.491 | **0.890** | 0.449 | 0.253 | 0.200 | 0.146 | 0.284 | 0.216 | 0.291 | 0.114 | 0.435 |
| Coordination and continuity of care | 0.408 | 0.315 | 0.457 | 0.467 | 0.430 | 0.423 | 0.440 | 0.341 | **0.675** | 0.544 | 0.517 | 0.399 | 0.492 | 0.507 | 0.274 | 0.379 | 0.557 |
| Patient safety | 0.513 | 0.420 | 0.568 | 0.551 | 0.520 | 0.521 | 0.323 | 0.216 | 0.642 | **0.820** | 0.566 | 0.415 | 0.592 | 0.610 | 0.392 | 0.391 | 0.477 |
| Patient information | 0.483 | 0.440 | 0.522 | 0.498 | 0.406 | 0.445 | 0.302 | 0.177 | 0.631 | 0.597 | **0.836** | 0.429 | 0.533 | 0.594 | 0.253 | 0.339 | 0.560 |
| Patient involvement in care | 0.379 | 0.358 | 0.538 | 0.463 | 0.365 | 0.412 | 0.149 | 0.114 | 0.454 | 0.435 | 0.416 | **0.757** | 0.417 | 0.516 | 0.313 | 0.375 | 0.681 |
| Involvement of family and friends | 0.604 | 0.583 | 0.573 | 0.562 | 0.511 | 0.601 | 0.389 | 0.322 | 0.714 | 0.720 | 0.644 | 0.514 | **0.962** | 0.681 | 0.529 | 0.477 | 0.323 |
| Patient empowerment | 0.673 | 0.595 | 0.753 | 0.691 | 0.679 | 0.688 | 0.366 | 0.203 | 0.749 | 0.755 | 0.700 | 0.629 | 0.674 | **0.909** | 0.489 | 0.594 | 0.424 |
| Physical support | 0.470 | 0.385 | 0.402 | 0.403 | 0.328 | 0.424 | 0.315 | 0.263 | 0.444 | 0.505 | 0.392 | 0.353 | 0.516 | 0.503 | **0.727** | 0.340 | 0.673 |
| Emotional support | 0.737 | 0.605 | 0.670 | 0.731 | 0.602 | 0.620 | 0.225 | 0.111 | 0.596 | 0.534 | 0.439 | 0.515 | 0.496 | 0.623 | 0.309 | **0.900** | 0.480 |

**Table 2:** Correlations between sum scores and model-based factor scores - inpatient sample

| Sum scores for specific dimensions / model-based factor scores | Essential characteristics of the clinicians | Clinician-patient relationship | Patient as a unique person | Biopsychosocial perspective | Clinician-patient communication | Integration of medical and non-medical | Teamwork and teambuilding | Access to care | Coordination and continuity of care | Patient safety | Patient information | Patient involvement in care | Involvement of family and friends | Patient empowerment | Physical support | Emotional support | General factor |
|---|---|---|---|---|---|---|---|---|---|---|---|---|---|---|---|---|---|
| Essential characteristics of the clinicians | **0.427** | 0.204 | 0.207 | 0.203 | 0.104 | 0.203 | 0.016 | -0.024 | 0.220 | 0.161 | 0.103 | 0.139 | 0.156 | 0.189 | 0.020 | 0.256 | 0.884 |
| Clinician-patient relationship | 0.356 | **0.654** | 0.380 | 0.365 | 0.334 | 0.300 | 0.158 | -0.070 | 0.286 | 0.277 | 0.238 | 0.270 | 0.240 | 0.329 | -0.036 | 0.398 | 0.751 |
| Patient as a unique person | 0.529 | 0.536 | **0.777** | 0.624 | 0.494 | 0.527 | 0.209 | -0.067 | 0.543 | 0.580 | 0.477 | 0.531 | 0.485 | 0.617 | 0.083 | 0.514 | 0.614 |
| Biopsychosocial perspective | 0.631 | 0.613 | 0.764 | **0.923** | 0.530 | 0.631 | 0.291 | -0.020 | 0.751 | 0.658 | 0.600 | 0.514 | 0.577 | 0.670 | 0.041 | 0.687 | 0.324 |
| Clinician-patient communication | 0.153 | 0.254 | 0.278 | 0.238 | **0.588** | 0.185 | 0.095 | -0.042 | 0.183 | 0.209 | 0.189 | 0.197 | 0.154 | 0.241 | -0.026 | 0.244 | 0.825 |
| Integration of medical and non-medical care | 0.638 | 0.514 | 0.636 | 0.636 | 0.395 | **0.950** | 0.268 | 0.082 | 0.697 | 0.585 | 0.405 | 0.495 | 0.562 | 0.666 | 0.103 | 0.629 | 0.331 |
| Teamwork and teambuilding | 0.024 | 0.095 | 0.108 | 0.119 | 0.096 | 0.124 | **0.538** | -0.084 | 0.177 | 0.157 | 0.143 | 0.057 | 0.163 | 0.134 | 0.044 | 0.102 | 0.800 |
| Access to care | -0.013 | -0.061 | 0.016 | 0.038 | 0.002 | 0.088 | -0.035 | **0.568** | 0.130 | 0.071 | 0.034 | 0.017 | 0.065 | 0.050 | 0.170 | 0.035 | 0.780 |
| Coordination and continuity of care | 0.337 | 0.244 | 0.328 | 0.361 | 0.198 | 0.346 | 0.201 | 0.071 | **0.529** | 0.359 | 0.338 | 0.273 | 0.344 | 0.331 | 0.015 | 0.309 | 0.783 |
| Patient safety | 0.345 | 0.287 | 0.490 | 0.455 | 0.300 | 0.411 | 0.254 | 0.022 | 0.528 | **0.737** | 0.437 | 0.335 | 0.461 | 0.523 | 0.097 | 0.314 | 0.597 |
| Patient information | 0.249 | 0.284 | 0.440 | 0.443 | 0.302 | 0.319 | 0.273 | 0.004 | 0.525 | 0.473 | **0.791** | 0.371 | 0.431 | 0.453 | 0.037 | 0.286 | 0.605 |
| Patient involvement in care | 0.320 | 0.328 | 0.480 | 0.381 | 0.291 | 0.390 | 0.086 | -0.010 | 0.422 | 0.394 | 0.378 | **0.753** | 0.366 | 0.383 | 0.019 | 0.307 | 0.663 |
| Involvement of family and friends | 0.431 | 0.355 | 0.524 | 0.526 | 0.310 | 0.535 | 0.303 | 0.024 | 0.659 | 0.583 | 0.506 | 0.434 | **0.908** | 0.512 | 0.123 | 0.424 | 0.385 |
| Patient empowerment | 0.552 | 0.526 | 0.707 | 0.648 | 0.468 | 0.598 | 0.309 | -0.019 | 0.644 | 0.709 | 0.576 | 0.484 | 0.540 | **0.889** | 0.083 | 0.556 | 0.406 |
| Physical support | 0.137 | 0.118 | 0.197 | 0.174 | 0.105 | 0.208 | 0.093 | 0.143 | 0.150 | 0.187 | 0.102 | 0.116 | 0.183 | 0.189 | **0.525** | 0.163 | 0.762 |
| Emotional support | 0.786 | 0.684 | 0.647 | 0.711 | 0.551 | 0.632 | 0.258 | 0.009 | 0.651 | 0.506 | 0.396 | 0.435 | 0.481 | 0.601 | 0.068 | **0.915** | 0.376 |
