## Appendix 8 Construct validity for "Through the patients’ eyes - Psychometric evaluation of the 64-item version of the Experienced Patient-Centeredness Questionnaire (EPAT-64)"

**Article:** Through the patients' eyes - Psychometric evaluation of the Experienced Patient-Centeredness Questionnaire (EPAT-64)

**Authors:** Eva Christalle, Stefan Zeh, Hannah Führes, Alica Schellhorn, Pola Hahlweg, Jördis Zill, Martin Härter, Carsten Bokemeyer, Jürgen Gallinat, Christoffer Gebhardt, Christina Magnussen, Volkmar Müller, Katharina Schmalstieg-Bahr, André Strahl, Levente Kriston, Isabelle Scholl

Table 1: Construct validity: Correlations between model-based factor scores and health status as well as satisfaction

| | **Outpatients** | | **Inpatients** | |
|---|---|---|---|---|
| **Dimension** | **Health status[1]** | **Satisfaction[2]** | **Health status[1]** | **Satisfaction[2]** |
| *Hypothesis about correlations* | *absolute <0.3* | *>0.5* | *absolute <0.3* | *>0.5* |
| Essential characteristics of the clinicians | -0.036 | 0.335 | -0.003 | 0.172 |
| Clinician-patient relationship | -0.045 | 0.299 | 0.013 | 0.129 |
| Patient as a unique person | 0.008 | 0.312 | -0.014 | 0.175 |
| Biopsychosocial perspective | -0.002 | 0.267 | -0.024 | 0.149 |
| Clinician-patient communication | -0.055 | 0.224 | 0.020 | 0.083 |
| Integration of medical and non-medical care | -0.016 | 0.276 | -0.065 | 0.154 |
| Teamwork and teambuilding | -0.058 | 0.237 | -0.108 | 0.143 |
| Access to care | -0.060 | 0.198 | -0.028 | -0.007 |
| Coordination and continuity of care | -0.055 | 0.339 | -0.059 | 0.193 |
| Patient safety | -0.034 | 0.320 | -0.014 | 0.176 |
| Patient information | -0.052 | 0.325 | -0.076 | 0.160 |
| Patient involvement in care | -0.022 | 0.264 | -0.019 | 0.152 |
| Involvement of family and friends | -0.060 | 0.321 | -0.044 | 0.164 |
| Patient empowerment | -0.040 | 0.324 | -0.057 | 0.167 |
| Physical support | -0.053 | 0.251 | -0.098 | 0.038 |
| Emotional support | -0.015 | 0.230 | -0.025 | 0.137 |
| General factor | -0.166 | 0.728 | -0.087 | 0.833 |

[1] General health status measured by first item of the German version of 12-Item Short Form Survey.[32]

[2] Treatment satisfaction measured by the German version of the 8-item Client Satisfaction Questionnaire (ZUF-8).[33]
